# Towards real-world molecular pathology diagnosis of cancer with cross-modal AI

**DOI:** 10.1101/2025.07.07.25330997

**Authors:** Xiaofei Wang, Yinyan Wang, Wanming Hu, Yupei Zhang, Xinke Zhang, Chen Lu, Jie Hu, Zeya Yan, Xing Liu, Hao Duan, Yonggao Mou, Tao Jiang, Mayen Briggs, Stephen Price, Chao Li

## Abstract

Accurate integration of histological and molecular features is central to modern cancer diagnostics, but it is often hampered by resource-intensive parallel workflows, limited tissues, and increased diagnostic complexity. We present CAMPaS (Cross-modal AI for Integrated Molecular Pathology Diagnosis and Stratification), a clinical AI prototype that addresses challenges in real-world translation of jointly predicting glioma histology, molecular markers, and WHO 2021 integrative diagnoses from hematoxylin and eosin-stained slides. Trained and validated on 3,367 patients (6,043 slides) across eight cohorts (six retrospective, two prospective), CAMPaS achieved high diagnostic performance (AUC 0.895-0.916 in training; 0.946-0.955 in prospective cohorts) and generalized robustly across diverse settings. Its interpretable cross-modal predictions aligned with histopathological annotations and genomic profiles, revealing biologically coherent features. CAMPaS identified histological features for molecular markers, and its clinical utility was validated for enhancing real-world clinical diagnostics. Crucially, CAMPaS stratifies prognosis and treatment response, offering a scalable and biologically grounded solution to accelerate precision oncology.

## Introduction

Accurate and timely cancer diagnosis is crucial for guiding treatment and improving patient outcomes. While pathology remains the diagnostic gold standard, traditional histopathology requires substantial expertise and is subject to interobserver variability^1^. Over the past decade, molecular profiling has revolutionized cancer diagnosis^2^. The 2021 WHO classification of central nervous system tumors reflects this paradigm shift, embedding key molecular markers into diagnostic criteria^3^. For instance, glioblastoma is no longer diagnosed solely on histological features^3–5^ but primarily diagnosed by *IDH* mutation status, even in the absence of high-grade histology. Nonetheless, histology remains essential. A category of grade 4 astrocytoma is newly defined by *IDH* mutated *1p/19q* non-co-deletion (CODEL), together with either *CDKN2A/B* homozygous deletion (HOMDEL) and/or histological features such as necrosis and microvascular proliferation.

While this integrative approach enhances diagnostic precision, it complicates routine clinical workflows^6^. First, molecular testing is costly and may be infeasible due to limited tissue availability^7^. Second, histological assessment and molecular testing follow separate, time-consuming pipelines, hindering timely integration for final diagnosis.

Third, incorporating molecular markers presents challenges for practitioners trained primarily in morphology, increasing the risk of misdiagnosis^8^. Particularly, the resource requirements make molecular diagnostics unavailable to patients in low- or middle-income countries.

Artificial Intelligence (AI) has enabled the direct prediction of molecular markers from hematoxylin and eosin (H&E)-stained slides, thereby bridging histological and molecular diagnostics. However, current AI approaches^10–13^ typically treat molecular and histological prediction as separate tasks, overlooking their biological interaction.

Additionally, most existing models are trained on small cohorts, limiting their performance. This limitation is particularly evident for histologically low-grade lesions with molecular aberrations that denote more aggressive clinical behavior.

Emerging studies demonstrate the feasibility of AI in modelling the complex relationships between histological and molecular features, offering the potential for more accurate, integrated diagnosis^13,14^. However, real-world challenges, including heterogeneous datasets and missing molecular data, may compromise model robustness. Although AI models can generate spatial maps and surrogate molecular signatures to support pathologists in interpreting tissue holistically^9,14^, few have rigorously validated their pathological coherence and interpretability at the histological and molecular levels. Moreover, rare models are prospectively evaluated across diverse clinical settings, limiting confidence in their safety and utility for clinical workflows.

Here, we introduce CAMPaS (Cross-modal AI for integrated Molecular Pathology Diagnosis and Stratification), a trustworthy AI framework tailored for real-world cancer diagnosis. To ensure *accuracy*, *robustness, interpretability*, and *clinical utility*, our **primary** objective is to develop our previously benchmarking-validated model that jointly predicts molecular markers and histology^13,14^ into a clinical prototype, specifically addressing real-world challenges. Our **secondary** objective is to rigorously validate the model using a large-scale dataset of 3,367 gliomas (6,043 slides) from eight cohorts, including two prospective cohorts, achieving strong accuracy and robust generalization. CAMPaS demonstrates interpretability at histological, molecular, and cross-modal levels, aligning with established knowledge and contributing to a deeper understanding of glioma. We prospectively validated its clinical utility in supporting pathologists’ diagnosis, demonstrating its potential to identify histological features indicative of molecular markers. Further, CAMPaS demonstrates clinical relevance by predicting patient prognosis and treatment benefit over conventional clinicopathological metrics.

The presented clinical prototype of AI for cancer diagnosis bridges conceptual model development with future regulatory-aligned clinical deployment, along the translation pathway^15^. Together, our results underscore the potential of a trustworthy AI framework, purposefully designed and rigorously validated, supporting precision oncology.

## Results

### 2.1 Study design

To develop a clinically translatable AI system for integrated cancer diagnosis, we extended our previously validated framework^13,14^ that jointly models molecular markers and histology. The proposed CAMPaS introduces a substantially enhanced architecture to address the key challenges of real-world deployment, including data heterogeneity, incomplete molecular labeling (Fig. 1a-b).

**Fig 1.**
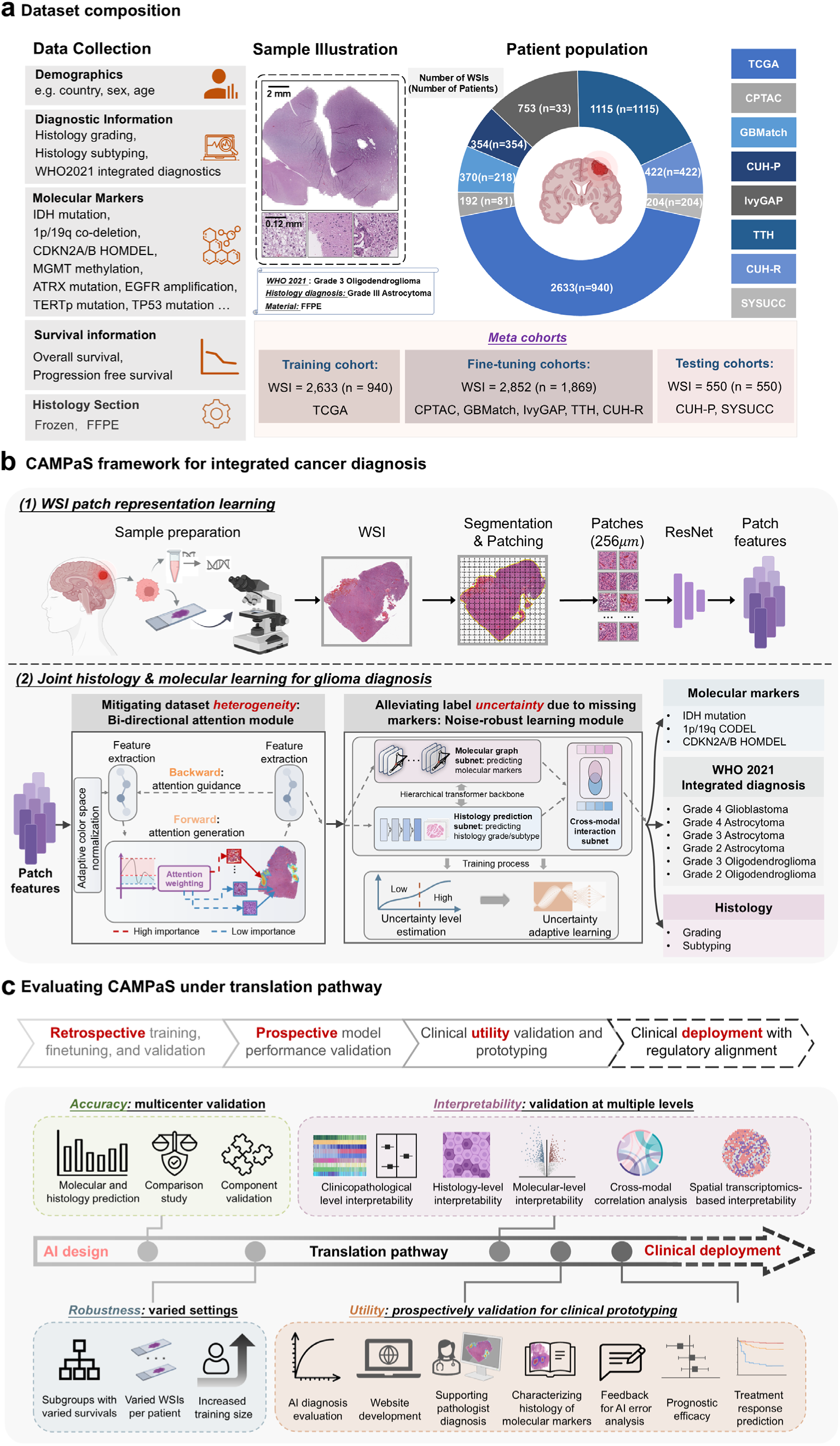
Data, model design, and validation. **a,** We include multi-modal data, including demographics, diagnosis, molecular markers from either immune-staining or sequencing, survival information, and whole slide images (WSIs). These data sources, whenever available, are aggregated from eight independent cohorts: TCGA, CPTAC, GBMatch, IvyGAP, Cambridge University Hospital (retrospective: CUH-R; prospective: CUH-P), Beijing Tian Tan Hospital (TTH), and Sun Yat-sen University Cancer Center (SYSUCC) (Tables S1). We first train the model on the large-scale TCGA cohort, and then fine-tune and validate it on other retrospective datasets (CPTAC, GBMatch, IvyGAP, CUH-R, and TTH cohorts). We further recruit CUH-P and SYSUCC cohorts for prospective validation. **b**, Illustration of the CAMPaS framework for integrated cancer diagnosis. The framework consists of two stages: (1) WSI patch representation learning and (2) joint histology and molecular learning for glioma diagnosis. In the **first** stage, tumor regions are segmented from WSIs, divided into 256 μm patches, and filtered to remove background and low-quality regions (e.g., inked or poorly stained areas). A pretrained ResNet^20^ is then utilized to extract patch-level features, with the last fully connected layers of the network excluded. See Supplementary section 1 for detailed WSI preprocessing and the patch representation learning process. In the **second** stage, CAMPaS jointly predicts histology grading/subtyping and molecular markers, finally integrated for glioma classification, with two specially designed modules, i.e., **bi-directional attention module** and **noise-robust learning module**, for mitigating challenges of dataset heterogeneity and label uncertainty in real-world datasets. Specifically, CAMPaS is a multi-task, multi-instance learning framework built upon a hierarchical vision transformer^21^ backbone. For each input WSI, the extracted WSI features from the first stage are fed into the bi-directional attention module to extract robust features. The **bi-directional attention module** dynamically weights the importance of WSI patches and therefore generates patch attention to identify and select critical regions of WSIs, guiding the feature extraction in a forward-backward paradigm. Additionally, this module incorporates an adaptive color-space normalization strategy to mitigate variability introduced by WSI processing procedures. Further, **the noise-robust learning module** refines the extracted features, enabling them to alleviate the label uncertainty caused by missing molecular markers. This is achieved by estimating the uncertainty of each WSI from the missing molecular information, followed by an adaptive learning process that accounts for the label noise resulting from missing information. The previously proposed modules, i.e., molecular graph, histology prediction, and cross-modal interaction subnets,^13,14^ are incorporated to model molecular markers, predict histology grading/subtyping, and achieve cross-modal interaction, respectively. Details of CAMPaS can be found in Methods and Extended Fig. 3. **c**, Evaluation of the CAMPaS under the translation pathway. Overall, our CAMPaS-based pipeline represents the clinical prototyping stage, encompassing prospective utility validation, bridging model development with future regulatory-aligned clinical deployment along the translation pathway^22,23^. Specifically, various validations are incorporated sequentially, including: (i) **accuracy**, supported by multicenter validation of molecular and histology prediction, comparison studies, and component validation; (ii) **robustness**, tested across subgroup analysis, varied WISs per patient, and increased training sizes; (iii) **interpretability**, assessed at multiple levels including clinicopathological, histological, molecular, cross-modal validation; (iv) **clinical utility**, through prospective validation in glioma diagnosis, web deployment, assistance for pathologists’ diagnosis, characterization of histological features of molecular markers, feedback for AI error analysis, prognostic efficacy, and treatment response prediction.

CAMPaS was developed and evaluated under the principles of trustworthy AI (Fig. 1c). The model was first trained and validated on six retrospective cohorts, with interpretability rigorously assessed at histological, molecular, and cross-modal levels. Further, to establish clinical utility, we conducted prospective validation in two independent cohorts, investigating the model’s performance in assisting diagnosis and prognosis. Finally, we implemented CAMPaS as a web-based diagnostic tool (http://translational-ai.online; Extended Fig. 1), supporting practical use cases such as intraoperative diagnosis, postoperative evaluation from limited tissue, and retrospective dataset re-annotation (we also released updated labels of the TCGA dataset as per the WHO 2021 criteria for public use).

### 2.2 Cohorts

We assembled a multi-institutional dataset of 3,367 patients with 6,043 WSIs. We collected patient demographics, diagnoses, molecular markers from either immune staining or sequencing, and survival across both retrospective and prospective cohorts representing diverse ethnical backgrounds (Table S1; Supplementary section 2).

- Retrospective cohorts: Six independent cohorts were used for model training and validation: four public cohorts of TCGA^16^ (n=1,122), CPTAC^17^ (n=99), IvyGAP^18^ (n=41), and GBMatch^19^ (n=225), and two in-house cohorts from Cambridge University Hospital (CUH-R, n=590, January 2019-October 2021), and Beijing Tiantan Hospital (TTH, n=1,121, February 2015-January 2018). After excluding patients with missing or poor-quality data to meet the minimum requirement, we retained 2,809 patients (5,485 WSIs).
- Prospective cohorts: We recruited 369 patients from CUH (CUH-P, November 2021-February 2024) and 210 patients from Sun Yat-sen University Cancer Center (SYSUCC, April 2022-April 2024). After exclusion, we analyzed 558 patients (558 WSIs) in total, all diagnosed under the WHO 2021 criteria.

WHO 2021 glioma diagnosis workflows and inclusion/exclusion criteria are detailed in Extended Fig. 2 and Fig. S1.

### 2.3 Model design

Aligned with the WHO 2021 adult glioma diagnosis criteria (Extended Fig. 2), CAMPaS (Fig. 1b, Extended Fig. 3; Methods) jointly predicted histology grades (i.e., Grade 2, 3, and 4), subtypes (i.e., oligodendroglioma, astrocytoma, and glioblastoma [GBM]), and key molecular markers (i.e., *IDH* mutation, *1p/19q* CODEL, and *CDKN2A/B* HOMDEL), enabling a fully integrated diagnosis from routine H&E slides.

CAMPaS builds upon our previously benchmarked and validated designs of a molecular graph subnet, a histology prediction subnet, and a cross-modal interaction subnet, enabling the capture of relationships among molecular markers and guiding cross-modal interactions. This study further validated the robustness of these designs.

Particularly, we substantially enhanced the architecture to directly address two major translational barriers. First, to mitigate WSI heterogeneity (e.g., variations in staining, cell density, and dispersion; Figs. S2-3; Supplementary section 3), we designed a bi-directional attention module to dynamically identify and select diagnostically salient WSI regions. In this module, an adaptive color-space normalization strategy was embedded to reduce the effects of WSI procedures.

Second, to address incomplete molecular markers (summary of missing markers of all cohorts in Table S2), a common limitation in real-world datasets, we designed a noise-robust learning module. This module estimates the uncertainty of each WSI using Shannon entropy and incorporates an uncertainty-adaptive learning strategy, down-weighting noisy or ambiguous labels during training to improve model resilience to label incompleteness.

By integrating these translational design elements, CAMPaS moves beyond prior cross-modal architectures to support robust learning on clinical cohorts.

### 2.4 Model performance on retrospective cohorts

#### 2.4.1 Accuracy analysis

We first trained CAMPaS on TCGA, achieving a mean AUC of 0.905 (95% CI: 0.895-0.916, *P*=0.041) for glioma classification, based on three-fold cross-validation with bootstrapping (Fig. 2a and Table S3). AUCs for the six WHO 2021 glioma subtypes ranged from 0.848 to 0.918 (all *P*<0.050; Table S4).

**Fig 2.**
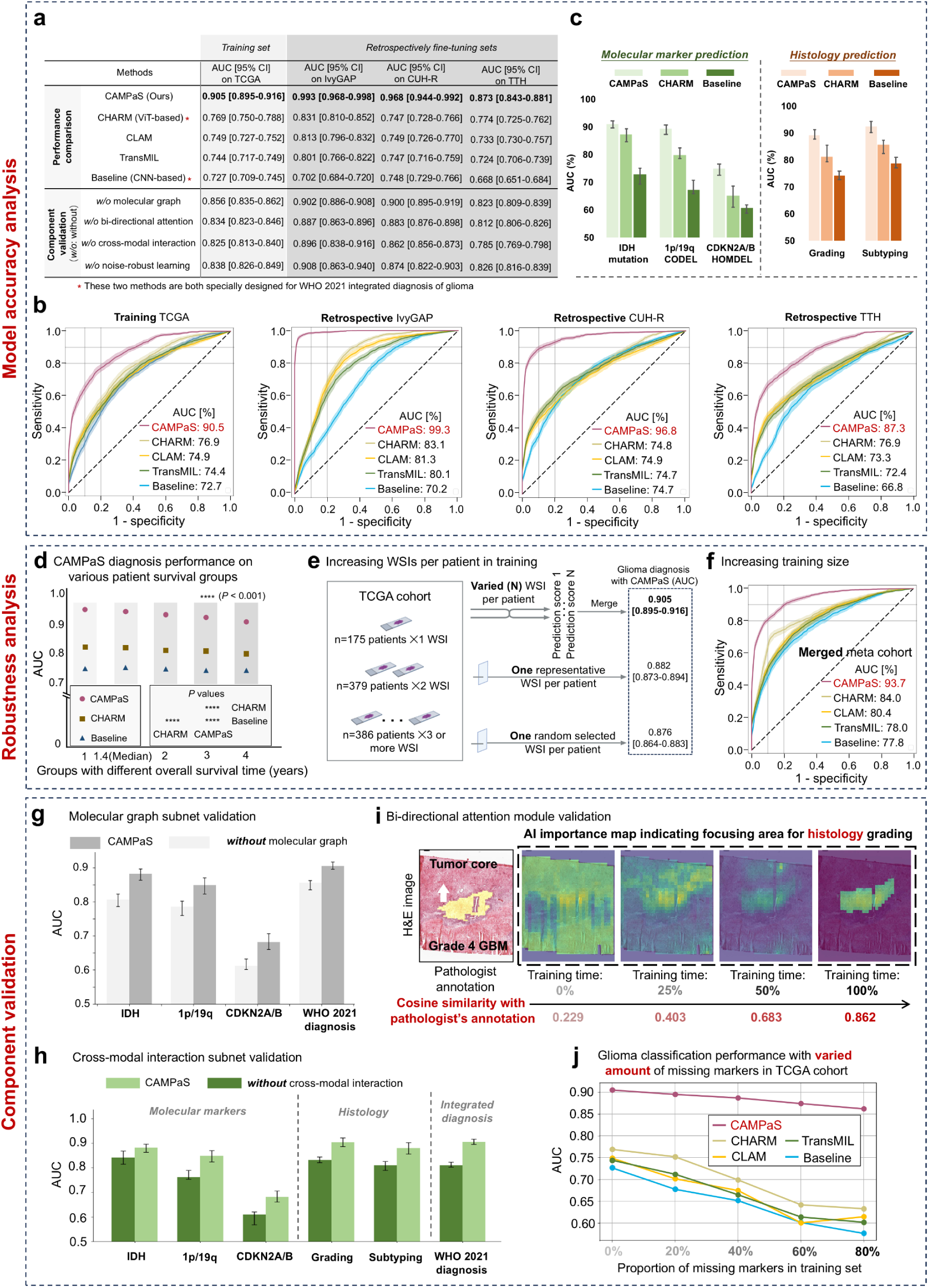
Validation on multicenter retrospective cohorts. **(a-c), Accuracy analysis. a,** *Top panel*: Comparison of CAMPaS against CHARM, CLAM, TransMIL, and a CNN-based baseline. *Bottom panel:* Ablation studies evaluating the contribution of the four key components proposed in CAMPaS. **b,** ROC curves of CAMPaS, CHARM, CLAM, TransMIL and the CNN baseline. **c,** Bar charts illustrating AUC values (with error bars indicating 95% CIs) for predicting molecular markers, histology grading, and subtyping, comparing CAMPaS with other methods. **(d-f), Robustness analysis. d,** Performances of CAMPaS and comparison methods in glioma classification in the TCGA cohort, measured by AUC, across patient subgroups stratified by overall survival (OS). Each subgroup includes patients with OS less than the specified threshold. CAMPaS consistently outperforms other methods on different survival subgroups, demonstrating robust performance. **e,** Comparing the default input setting, i.e., using multiple WSIs per patient, with alternative input strategies: (i) one randomly selected WSI, and (ii) the most representative (i.e., with the smallest prediction uncertainty) WSI on the TCGA cohort (n = 940). **f,** ROC curves on the merged cohort comparing CAMPaS and competing methods, highlighting the superiority of CAMPaS trained on all cohorts over its TCGA-training-only variant and other methods, further demonstrating its robustness. **(g-j), Model component validation. g,** Evaluation of the proposed molecular graph subnet on molecular marker predictions and final diagnosis, demonstrating its effectiveness in modeling correlations of molecular markers to enhance integrated diagnosis. **h,** Evaluation of the proposed cross-modal interaction subnet. Removing this module leads to substantial performance drops across molecular marker prediction, histology grading/subtyping, and integrated diagnosis, highlighting its critical role in integrating multimodal information for improved diagnostic accuracy. **i,** Assessment of the bi-directional attention module. As training progresses, AI-generated importance maps for histology grading become progressively denoised and gradually focus on the tumor core, indicating improved patch selection and feature robustness. **j,** Glioma classification performance under increasing proportions of missing molecular markers in the TCGA training set. CAMPaS consistently outperforms other methods (CHARM, TransMIL, CLAM, and Baseline) across all levels of missingness, demonstrating the superiority of the proposed noise-robust learning module in real-world scenarios of missing molecular markers.

To enhance generalizability on heterogeneous datasets, we fine-tuned CAMPaS on five additional retrospective cohorts, observing high and consistent performance across diverse datasets (Table S3): CPTAC (AUC: 0.976, 95%CI: 0.952-1.000, *P*=0.022), GBMatch (AUC: 0.981, 95%CI: 0.956-0.992, *P*=0.013), IvyGAP (AUC: 0.993, 95%CI: 0.968-0.998, *P*=0.018), TTH (AUC: 0.873, 95%CI: 0.843-0.881, *P*=0.040), and CUH-R (AUC: 0.968, 95%CI: 0.944-0.992, *P*=0.016).

CAMPaS was benchmarked against four comparison methods: CHARM^11^ (a hierarchical vision transformer^21^ [ViT]-based model as the SOTA comparator); CLAM^24^ and TransMIL^25^ (widely used multi-instance learning [MIL] methods), and a CNN-based baseline (proposed for glioma classification^9^). On TCGA, CAMPaS outperformed all comparison methods, with a ≥13.6% (*P* < 0.001) AUC improvement, while a ≥6.1% (*P* < 0.001) improvement was observed across all five retrospective fine-tuning cohorts (Table S3). ROC curves (Fig. 2b and Fig. S4; all *P*<0.001; Delong test) and forest plots (Fig. S5) confirmed its superior performance. Notably, CAMPaS achieved an AUC of 0.907 for the newly classified Grade 4 astrocytoma, substantially outperforming CHARM (AUC 0.608) and the CNN-based baseline (AUC 0.486), highlighting its strong generalizability to rare but clinically important subtypes.

CAMPaS also performed strongly on auxiliary tasks of predicting histology grades/subtypes and molecular markers (Fig. 2c; Table S5; Supplementary section 5). For example, in TCGA, it achieved AUCs of 0.911 (*IDH* mutations, 95% CI: 0.899-0.925, *P*=0.015), 0.893 (*1p/19q* CODEL, 95% CI: 0.879-0.913, *P*=0.017) and 0.749 (*CDKN2A/B* HOMDEL, 95% CI: 0.733-0.771, *P*=0.022), consistently outperforming other methods (*P*<0.001).

#### 2.4.2 Robustness analysis

To evaluate robustness, we performed subgroup analyses based on sex, histology subtype and grade, section type, survival time, and diagnostic grade shift^1^ in both training (TCGA) and retrospective validation cohorts (TTH, CPTAC, GBMatch, IvyGAP) (Fig. 2d; Fig. S6; Tables S6-11; Supplementary section 6). For example, the model remained comparable between the TCGA subsets with consistent grading (AUC 0.913) and shifted grading (AUC 0.896). This stability shows that CAMPaS can adapt to both histology-dominant and integrated diagnoses under different WHO criteria, demonstrating robustness to criteria variations. Additionally, CAMPaS demonstrated high performance on both frozen (AUC 0.900) and FFPE (AUC 0.940) sections in TCGA, indicating robustness in both intraoperative and postoperative diagnosis scenarios.

We also tested model robustness on TCGA patients with multiple WSIs (n=765, 81.4%). Using a single random slide per patient yielded an AUC of 0.876, which improved slightly to 0.882 when the most representative slide was used (Methods). Notably, aggregating predictions across all WSIs (i.e., default setting in CAMPaS) improved AUC to 0.905 (Fig. 2e), demonstrating that increasing data diversity enhances model robustness.

Finally, we trained CAMPaS on the merged dataset covering all six cohorts to leverage the advantages of scale and diversity for enhancing model performance and generalization. CAMPaS outperformed all compared methods on all metrics, with an AUC of 0.937 (95% CI: 0.929-0.953, *P*=0.024), surpassing CHARM (AUC 0.912), CLAM (AUC 0.804), TransMIL (AUC 0.780), CNN baseline (AUC 0.840), and CAMPaS variant trained solely on TCGA (AUC 0.905) (Fig. 2f; Table S12). This indicates that expanding the training size enhances model robustness and generalization.

#### 2.4.3 Component validation

We validated the effectiveness of key components in CAMPaS through ablation studies, removing each one at a time (Fig. 2a, g-k). Removing the molecular graph subnet and cross-modal interaction subnet resulted in significant performance drops (Fig. 2g-h), with AUC decreasing from 0.968 to 0.900 (*P*=0.012) and 0.862 (*P*=0.024) in CUH-R, respectively, highlighting their roles in modeling molecular correlations and histology-to-molecular interactions.

Removing the bi-directional attention module caused a decrease in AUC on TCGA from 0.905 (95% CI: 0.895-0.916, P=0.045) to 0.834 (95% CI: 0.823-0.846, P=0.047) (Fig. 2a). During training, AI showed a gradual focus on the tumor core, finally aligning closely with expert annotations (Fig. 2i), confirming the role of the bi-directional attention module in identifying key regions with robust features for accurate prediction.

Furthermore, removing the noise-robust learning module consistently decreased performance across all cohorts, such as IvyGAP (AUC from 0.993 to 0.908) and CUH-R (AUC from 0.968 to 0.874) (Fig. 2a). Additionally, when trained with increasing levels of missing molecular markers (from 0% to 80% in TCGA), CAMPaS showed only a slight decline (AUC from 0.905 to 0.862), while consistently outperforming all comparison methods (P < 0.001; Fig. 2j). This indicates that this learning strategy effectively mitigates label noise by down-weighting noisy or ambiguous samples, thus preventing overfitting caused by label uncertainty and enhancing model stability under real-world conditions.

### 2.5 Model interpretability analysis

#### 2.5.1 AI-derived features are associated with clinicopathological signatures

To assess the interpretability of CAMPaS, we analyzed correlations between AI-derived features and clinicopathological signatures. Following prior work^26,27^, we stratified TCGA patients into low (n = 305), intermediate (n = 102), and high (n = 533) malignancy groups using agglomerative hierarchical clustering^28^ based on AI-predicted malignancy scores (Methods; Extended Fig. 4).

Multiple linear regression (Fig. 3a-b; Table S13) demonstrated that higher AI-predicted malignancy was positively linked to known pathology signatures indicating poor prognosis, including necrosis, angiogenesis, and IDH wildtype. These connections were also observed in the prospective SYSUCC and CUH-P cohorts (Fig. S7), further supporting the clinical relevance and interpretability of CAMPaS predictions.

**Fig 3.**
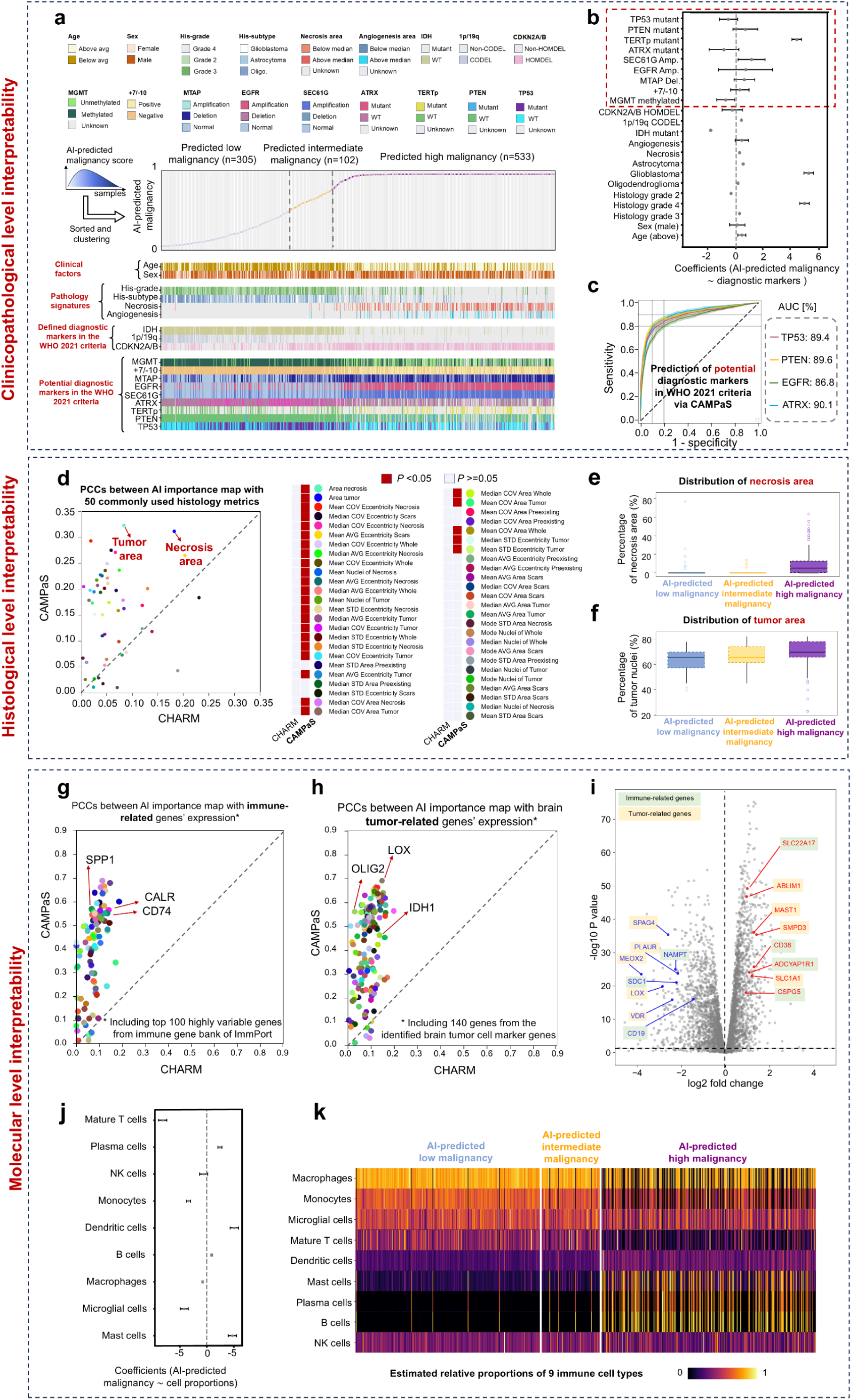
Model interpretability analysis. **(a-c) Clinicopathological level interpretability. a,** Integrated visualization of AI-predicted malignancy scores and clinical factors, pathology features, and molecular markers in the TCGA cohort (n = 940). The top panel shows the AI-predicted malignancy score curve across patients, stratified into three subtypes using agglomerative clustering. The bottom panel displays heatmaps of key clinical, histological, and molecular features, including defined diagnostic markers and potential markers as per the WHO 2021 criteria. *+7/-10* denotes the presence of concomitant gain of chromosome 7 and loss of chromosome 10. **b,** Associations of AI-predicted malignancy scores with clinical factors, pathology features, and molecular markers, using multiple linear regression with the malignancy scores as the dependent variable. Data are presented as the coefficients of the linear regression and 95% CIs (n = 940). Amp.: amplification; Del.: deletion. **c,** ROC curves of predicting potential diagnostic markers (including *EGFR* amplification, *ATRX* mutation, *TP53* mutation, and *PTEN* mutation) that are not yet included in WHO 2021, using CAMPaS pretrained on the WHO 2021 glioma diagnosis task in the TTH cohort. The prediction performance demonstrates the potential of extending CAMPaS to future WHO criteria, incorporating additional diagnostic markers. **(d-f) Histological level interpretability. d,** Comparison of CAMPaS with CHARM (ViT-based model) towards histological interpretability. Scatter plots display Pearson correlation coefficients (PCCs) comparing the correlations between 50 expert-annotated histology metrics and AI importance maps in GBMatch. Each colored dot represents a single histological feature, with notable examples such as Tumor area and the Necrosis area. Histology metrics significantly correlated with AI importance maps (*P* < 0.05) are indicated by red squares in the adjacent legend. **e-f,** Distribution of histological features (**e,** percentage of necrosis area and **f,** percentage of tumor nuclei) across CAMPaS-predicted malignancy subtypes in the TCGA cohort. The box plots are defined by the center tick as the median value, the lower and upper parts of the box Q1 and Q3 quartiles, respectively, and the bounds of whiskers are (Q1 − 1.5 × IQR, Q3 + 1.5 × IQR). Any outlier points beyond the whiskers are displayed with point marks. **(g-k) Molecular level interpretability. g-h,** Correlation analysis between the AI importance maps of molecular markers and expression levels of 140 tumor-related (well-established glioma marker genes from literature^3,31^) (**g**) and 100 immune-related (top 100 highly variable genes from the Immune Gene Bank of ImmPort^32^) (**h**) genes in TCGA. These plots indicate that the importance maps of CAMPaS more strongly associate with gene expression signatures (both immune and tumor-related) compared to those of CHARM, supporting its enhanced molecular-level interpretability. **i,** Differential expression analysis between CAMPaS predicted high malignancy versus low malignancy in TCGA. It shows that tumors predicted as high or low malignancy by CAMPaS have distinct gene expression patterns, especially in immune- and tumor-related genes. Statistical significance was accepted < 0.050 (i.e., above the horizontal dashed line). **j-k,** Immune landscape across AI-predicted malignancy in TCGA gliomas. **j,** Multiple single linear regressions reveal associations between AI-predicted malignancy score and immune cell abundances. Normalized regression coefficients (log₂ fold change with 95% CI) indicate that dendritic cells, mast cells, plasma cells, and B cells are positively associated with AI-predicted malignancy scores, while mature T cells, microglial cells, monocytes, and macrophages show negative associations. These findings are consistent with prior knowledge^33–35^, suggesting that as malignancy increases, the immune environment becomes more suppressive and less cytotoxic, potentially facilitating immune evasion and tumor progression. **k,** Hierarchical clustering heatmap of abundance of 9 common human immune cells^36^ for TCGA patients (n = 940), ordered by three subtypes in terms of AI-predicted malignancy. Notably, mature T cells, microglial cells, monocytes, and macrophages are abundant in tumors with low and intermediate AI-predicted malignancy scores, whereas dendritic cells, mast cells, plasma cells, and B cells are more enriched in tumors with high AI-predicted malignancy scores. These patterns are consistent with the quantitative findings in (**j**).

We examined associations between AI-predicted malignancy scores with established diagnostic markers not yet included in the WHO 2021 classification (e.g., mutation status of *ATRX*, *TERTp*, *PTEN*, *TP53*), as well as other reported prognostic markers (e.g., *MGMT* promoter methylation, *MTAP* deletion, *SEC61G* amplification) (Fig. 3a-b). For instance, *MGMT* promoter methylation (*P*=0.023) was linked to lower AI-predicted malignancy, aligning with their prevalence in *IDH*-mutated astrocytoma. *TERTp* mutation (*P*=2.22×10⁻⁹) was associated with higher scores, likely reflecting its co-occurrence with *IDH* wildtype in glioblastoma. Most markers are biologically related to *IDH* mutations, used in training CAMPaS, suggesting the model captures relevant molecular features beyond those directly supervised markers, based on tissue morphology.

Furthermore, we fine-tuned CAMPaS to predict established diagnostic markers not yet included in the WHO 2021 classification, such as *ATRX* mutation (AUC=0.832), *PTEN* mutation (AUC=0.865), *TP53* mutation (AUC=0.856), and *EGFR* amplification (AUC=0.890), in TTH (Fig. 3c). The accurate prediction highlights the transferability and potential usefulness of CAMPaS in future WHO criteria with additional diagnostic markers.

We expanded the analysis using the top 5% most prevalent molecular alterations from cBioPortal^29^ (Fig. S8). Several markers showed significant associations with AI-predicted malignancy. For instance, alterations in the well-established RTK/RAS/PI3K pathway^30^ (e.g., *PIK3CA*, *NF1*, *RB1*) were enriched in the high malignancy subgroup, consistent with their roles in glioma aggressiveness, suggesting model interpretability. Beyond established pathways, in the intermediate malignancy subgroup, we also observed enrichment of *CIC* and *FUBP1* mutations, which are involved in the SWI/SNF (SWItch/Sucrose Non-Fermentable) complex and histone acetylation/deacetylation pathways, indicating a potential role in glioma invasion and progression. This finding suggests that CAMPaS may identify potential diagnostic or prognostic markers, primarily attributed to its accurate prediction of hallmark markers, such as *IDH* mutations, and robust features learned from modelling molecular marker relationships.

#### 2.5.2 AI-derived features correlated with histological features

To validate histological relevance, we extracted AI importance maps^2^ for histology grading and calculated Pearson’s correlation coefficients (PCCs) between the AI importance maps and 50 pathologist-annotated histological features in the GBMatch cohort (Fig. 3d; Fig. S9). Twenty-seven features showed significant correlations (*P*<0.05), including established markers such as tumor area (*P*=0.0013) and necrosis area (*P*=0.0025) (Fig. 3d). Compared to other models, CAMPaS consistently demonstrated stronger correlations with histological metrics, using its top important (from top 50% to 5%) features^3^ (Fig. S9), showing alignment with expert annotations.

We further validated these findings in TCGA by comparing AI-defined malignancy subtypes. The high-malignancy group consistently displayed higher necrosis area and more tumor nuclei than the intermediate and low subtypes (Fig. 3e-f, Fig. S10), demonstrating the ability of CAMPaS to capture biologically and clinically relevant histological patterns.

#### 2.5.3 AI-derived features reflected tumor- and immune-related transcriptions

To assess model interpretability at the molecular level, we next analyzed transcriptional correlates of AI-predicted malignancy in TCGA (Fig. 3g-h, Figs. S11-S12), focusing on tumor-related and immune-related signature genes. CAMPaS consistently showed stronger correlations with gene expressions over the comparison models across both gene sets. Alongside the known genes such as *IDH* (PCC=0.45, *P*=0.006), *OLIG2* (PCC=0.56, *P*=0.003), other significant tumor-related genes included LOX (PCC=0.69, *P*=0.002), involved in extracellular matrix (ECM) remodeling and tumor invasion. For immune-related genes, significant correlations with gene expressions were observed for *SPP1* (PCC=0.56, *P*=0.002), *CALR* (PCC=0.56, *P*=0.001), and *CD74* (PCC=0.55, *P*=0.005), all known as immune-modulatory genes linked to tumor immune evasion. These results indicate the ability of CAMPaS to uncover potential markers of glioma invasiveness based on morphological features.

Differential expression analysis showed that high AI-predicted malignancy was associated with upregulated genes (Fig. 3i), such as *ABLIM1*, *MAST1*, and *SLC1A1*, linked to tumor invasion and progression. Conversely, the AI-predicted low-malignancy group showed enriched genes associated with hormone signaling, such as *VDR*, involved in Vitamin D signaling and anti-proliferative effects. Gene ontology enrichment analysis showed that these immune-related genes were involved in antigen presentation and immune response, while tumor-related genes were related to tissue remodeling, key features of tumor growth and invasion (Fig. S13).

Further, we employed BayesPrism^37^ to estimate the relative proportions of 9 common glioma immune cells with single-cell RNA sequencing^36^ as reference and analyzed their associations with AI-predicted malignancy scores (Fig. 3j-k, Table S14; Methods).

Multiple linear regression revealed that higher malignancy scores were positively correlated with dendritic cells (*P*=1.18×10^-9^), mast cells (*P*=1.03×10^-7^), plasma cells (*P*=4.44×10^-7^), and B cells (*P*=0.0082), indicating that higher AI-predicted malignancy is associated with elevated infiltration by immunosuppressive and inflammatory cells, suggesting the potential to guide patient selection for immune-targeted interventions.

These results collectively suggest that CAMPaS captures transcriptional and immunological patterns associated with tumor aggressiveness, demonstrating interpretability at the molecular level.

#### 2.5.4 CAMPaS showed interpretability across histological and molecular levels

To validate the cross-modal interpretability of CAMPaS, we generated AI importance maps from histology grade, subtype, and molecular marker predictions. Spatial correlation analyses demonstrated strong spatial alignment of these maps (Fig. 4a-b,d). For instance, AI importance maps for *IDH* wildtype spatially overlapped with those for Grade III and IV (PCC=0.495), while maps for *1p/19q* co-deletion closely aligned with those for oligodendroglioma (PCC=0.485). Further, compared to *1p/19q* CODEL and *CDKN2A/B* HOMDEL, the AI importance map for *IDH* wildtype showed strong correlations with pathologists-annotated necrosis and angiogenesis scores, aligning with the biological priors of high-grade gliomas^38,39^ (Fig. 4c). These spatial co-occurrence patterns suggest that the cross-modal interaction subnet can effectively model the relationships between histological and molecular features, enhancing cross-modal interpretability.

**Fig 4.**
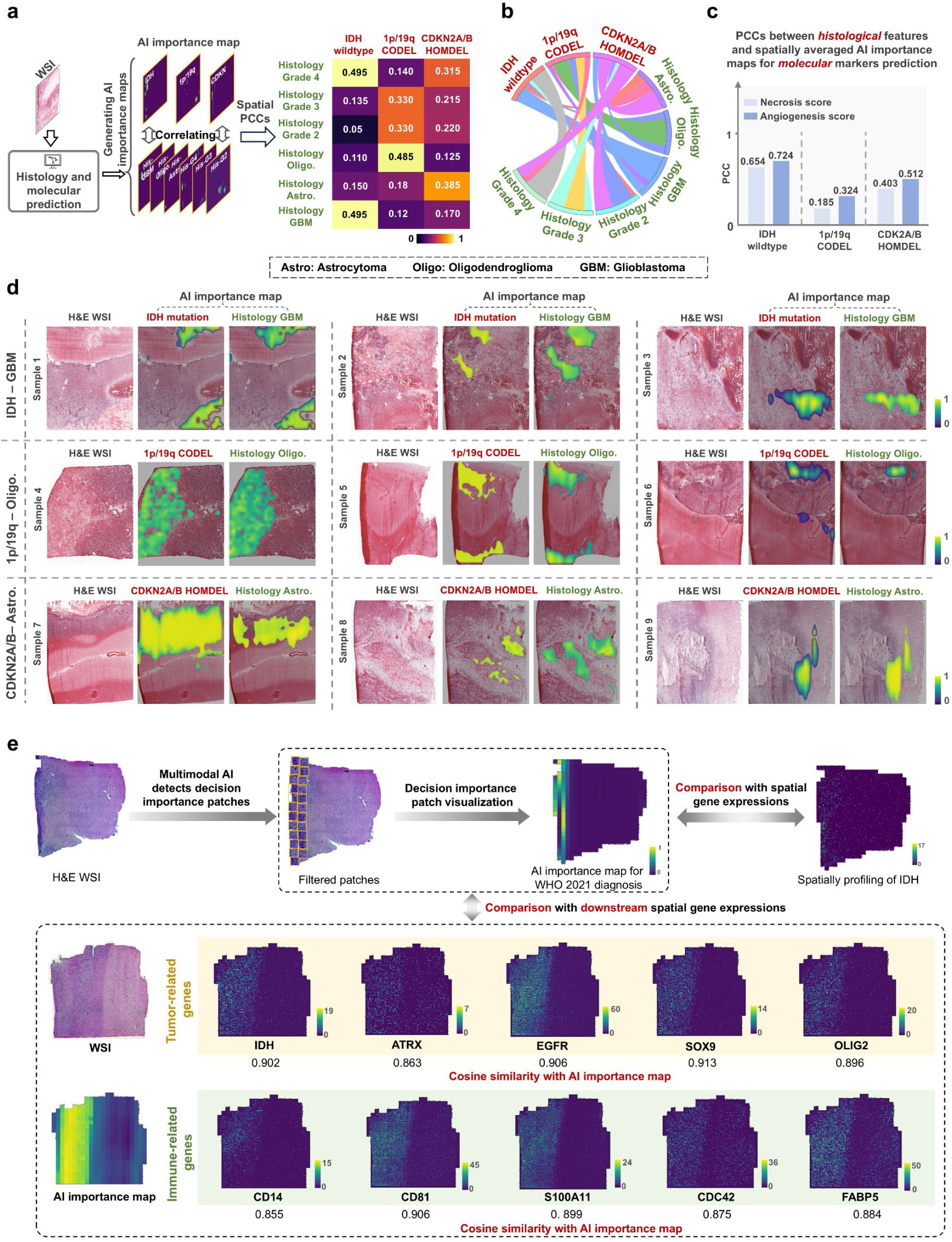
Cross-modal interpretability. **a,** Heatmaps showing the pairwise spatial Pearson correlation coefficients (PCCs) between AI importance maps of molecular markers prediction (i.e., *IDH* mutation, *1p/19q* CODEL, *CDKN2A/B* HOMDEL) and histological predictions (i.e., grades and subtypes). Oligo.: oligodendroglioma; Astro.: astrocytoma. **b,** Chord diagram visualizing the relationships in (**a**), highlighting the connections between AI-predicted molecular markers and histology features. The consistency with prior knowledge, e.g., histology subtype GBM and *IDH* wildtype, indicates the cross-modal interpretability of CAMPaS. **c,** Bar plots showing PCCs between histological features (pathologists-annotated angiogenesis score, necrosis score) and spatially averaged AI importance maps for predicting different molecular markers. For the AI importance map of *IDH* wildtype, correlations with necrosis and angiogenesis are both high, consistent with the aggressive and heterogeneous nature of high-grade glioma. In contrast, for the AI importance map of *1p/19q* CODEL, correlations with both metrics are low, reflecting the less aggressive, infiltrative tumor biology, further supporting the biological validity of the cross-modal spatial associations. **d,** Examples with strong spatial correlations observed between AI importance maps for molecular and histology prediction: spatial features associated with *IDH* wildtype status tend to align with those of histology GBM, while spatial features of *1p/19q* CODEL align with histology oligodendroglioma, and spatial features of CDKN2A/B HOMDEL align with histology astrocytoma (quantified in **a**). These paired maps exhibit consistent spatial patterns, reflecting known associations and supporting the model’s cross-modal interpretability. **e,** Spatial transcriptomics-based cross-modal interpretability validation. The upper panel illustrates the workflow for validating AI importance maps using spatial transcriptomics data^40^ (See Methods for data processing details): the model identifies decision-relevant patches, whose spatial correlations with spatial transcriptomic profiles are measured by cosine similarity. The lower panel shows spatial expression maps of selected genes. Tumor-related genes (*IDH*, *ATRX*, *EGFR*, *SOX9*, *OLIG2*) exhibit high spatial similarity (cosine similarity > 0.86) with AI importance maps, supporting that the model focuses on biologically meaningful tumor regions. These genes are key drivers of tumorigenesis and are common alterations defined by the WHO 2021 criteria. Similarly, immune-related genes (*CD14*, *CD81*, *S100A11*, *CDC42*, *FABP5*), associated with macrophage activation, immune signaling, and tumor–immune interactions, also show strong spatial concordance with AI-focused regions. This highlights the ability of CAMPaS to capture not only tumor-intrinsic features but also immune microenvironmental signals, suggesting the biological interpretability of AI decisions at both tumor and immune levels.

We further employed sequencing-based spatial transcriptomics and observed that decision-relevant spatial regions, highlighted by CAMPaS, showed strong spatial concordance with the expressions of tumor-related genes (e.g., *ATRX*, *EGFR*, *SOX9*, *OLIG2*) and immune-related genes (e.g., *CD14*, *CD81*, *S100A11*, *CDC42*, *FABP5*) (Fig. 4e; Fig. S14). Particularly, AI-highlighted regions exhibited strong spatial alignment with the expressions of many downstream genes of *IDH*, with cosine similarity scores ranging from 0.855 (*CD14*) to 0.913 (*SOX9*).

These findings highlight the ability of CAMPaS to produce spatially coherent and biologically relevant predictions across histological and molecular levels, reinforcing its interpretability and translational potential.

### 2.6 Prospective validation of clinical utility

#### 2.6.1 AI assistance improved diagnostic accuracy and reduced diagnostic discrepancy

We assessed the prospective generalizability of CAMPaS through zero-shot validation, training only on TCGA and testing on two independent prospective cohorts of CUH-P (n=354) and SYSUCC (n=204) without fine-tuning (Fig. 5a-b). Despite differences in tissue processing, scanner vendors, and patient demographics and ethnicity, CAMPaS demonstrated robust diagnostic performance (AUC: 0.946 for CUH-P, 0.955 for SYSUCC), largely attributed to its bi-directional attention module for handling WSI heterogeneity. Additionally, following prior work^41,42^, we quantified shifts in latent feature distributions for molecular marker prediction using Jensen-Shannon divergence (JSD). Prospective-to-retrospective JSD values (all<0.1) were comparable or lower than average retrospective-to-retrospective values (Fig. 5c), highlighting the stability and prospective generalizability of CAMPaS.

**Fig 5.**
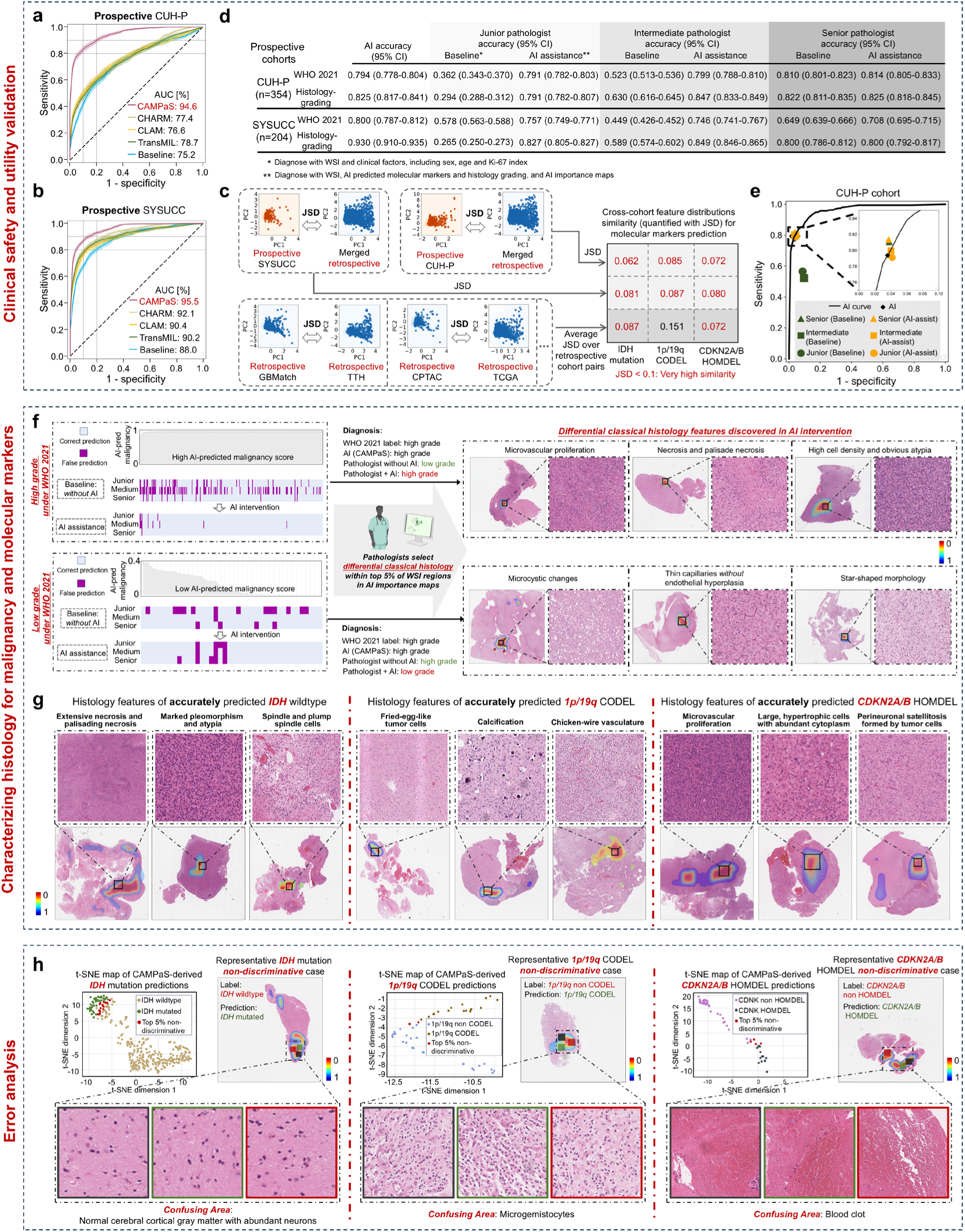
Prospective clinical utility validation of CAMPaS in assisting diagnosis. **(a-b),** ROC curves of CAMPaS, CHARM, CLAM, TransMIL, and the CNN baseline demonstrate that CAMPaS consistently achieves superior classification performance in the prospective CUH-P (**a**) and SYSUCC (**b**) cohorts. **c,** Jensen-Shannon divergence (JSD^41,42^) measuring how latent feature space of CAMPaS (derived via principal component analysis) for molecular markers prediction shifts from one cohort to another, including prospective SYSUCC/CUH-P to the merged retrospective cohorts (TCGA, CAPTAC, IvyGAP, GBMatch, TTH, and CUH-R), and the averaged pair-wise comparison among retrospective cohorts. Across all three markers of *IDH* mutation, *1p/19q* CODEL and *CDKN2A/B* HOMDEL, the prospective-retrospective JSD values are comparable to the retrospective mean, indicating that CAMPaS extracts feature representations in prospective cohorts that closely match those in retrospective data, underscoring its strong translational generalization capability. PC: principal component. **d,** Diagnostic accuracy of AI, pathologists, and AI-assisted pathologists across prospective cohorts. Accuracy for WHO 2021 classification and histology grading is reported for junior, intermediate, and senior pathologists in the CUH-P (n = 354) and SYSUCC (n = 203) cohorts. Across all experience levels, AI-assisted diagnosis significantly improves accuracy, notably for junior and intermediate pathologists, whose performance approaches that of senior pathologists and standalone AI. This highlights the effectiveness of CAMPaS in supporting clinical decision-making, particularly for less experienced pathologists. **Baseline:** using histology images and clinical factors. **AI assistance:** supported by AI-predicted histology grading, molecular markers, and AI importance maps. **e,** ROC curves illustrate the standalone AI performance (black line and diamond) alongside the sensitivity and specificity of senior, intermediate, and junior pathologists, both before and after receiving AI assistance, in the CUH-P cohort. **f,** Impact of AI intervention on diagnostic accuracy in glioma grading. Left panels illustrate changes in pathologists’ grading accuracy before (baseline) and after AI assistance in CUH-P. With AI assistance, junior and intermediate pathologists notably improved grading consistency, reducing misclassifications. Right panels demonstrate representative histological features discovered from AI importance maps as discriminative by pathologists. **g,** Representative histological features identified by CAMPaS in accurately predicting key molecular markers. For *IDH*-wildtype gliomas, CAMPaS highlights histological features such as extensive necrosis and palisading necrosis, marked pleomorphism and atypia, and spindle and plump spindle cells. For tumors with *1p/19q* CODEL, indicative features include round nuclei with perinuclear halos, fired-egg-like tumor cells, calcification, and chicken-wire vasculature. Meanwhile, for gliomas with *CDKN2A/B* HOMDEL, representative features are microvascular proliferation, large and hypertrophic cells with abundant cytoplasm, and perineuronal satellitosis formed by tumor cells. Notably, CAMPaS captures not only established histological hallmarks but also reveals potential indicators of molecular status, e.g., the co-occurrence of perivascular lymphocytic cuffing and classic high-grade features such as necrosis in IDH-wildtype tumors, offering potential insights into histology-molecular associations. **h,** Characterization of histological confounders associated with CAMPaS misclassification cases in molecular predictions. t-SNE plots highlight misclassified cases with non-discriminative features across three molecular alterations: *IDH* mutation, *1p/19q* CODEL, and *CDKN2A/B* HOMDEL. Pathological review revealed representative histological confounders associated with misclassification, including normal cerebral cortical gray matter with abundant neurons for misclassified *IDH* mutation, microgemistocytes for misclassified *1p/19q* CODEL, and blood clot regions for misclassified *CDKN2A/B* HOMDEL. These findings could potentially be used for guiding CAMPaS towards better performance and clinical interpretability through feedback integration of confounding histological features.

To assess clinical utility in diagnosis, we conducted a two-stage evaluation involving three neuropathologists (junior, intermediate, senior) using the WHO 2021 integrated criteria with both CUH-P and SYSUCC on our website (Extended Fig. 5). In stage I after surgery with tumor samples routinely obtained and processed, each pathologist reviewed anonymized cases independently based on WSIs and patient basic demographics. In stage II, they reviewed the above anonymized cases shuffled by algorithm. They were aided by CAMPaS prediction, including AI importance maps.

The diagnostic accuracy of both the pathologists and the AI system was assessed against the final WHO diagnosis confirmed by molecular testing. The results show that AI assistance significantly enhanced diagnostic accuracy across all pathologists and markedly reduced inter-pathologist discrepancy, highlighting its clinical utility.

Particularly, junior’s accuracy improved from 0.362 to 0.791 in CUH-P, and from 0.578 to 0.757 in SYSUCC. The intermediate improved similarly from 0.523 to 0.799 (CUH-P), and from 0.449 to 0.786 (SYSUCC). The senior pathologist, who already performed well, showed modest improvements, e.g., from 0.819 to 0.841 in CUH-P (Fig. 5d-e; Fig. S15). Similar gains were observed in AI-assisted histology grading (Fig. 5d). Notably, diagnostic performance across all pathologists became more consistent following AI assistance. Finally, CAMPaS achieved an average system usability scale (SUS) score of 76.8, outperforming previously reported standard benchmark^43^ (SUS=68) (Tables. S15-16).

The visual interpretability of CAMPaS guided pathologists toward regions most predictive of glioma malignancy (Fig. 5f). Specifically, in AI-corrected pathologist diagnoses in CUH-P, the top 5% of CAMPaS-highlighted regions corresponded to well-established markers of high malignancy, such as microvascular proliferation, high cell density, and obvious atypia. Meanwhile, regions indicative of lower malignancy were often characterized by microcystic changes, thin capillaries without endothelial hyperplasia, and star-shaped morphology.

Further analysis revealed distinct histological patterns for accurately predicted key molecular markers in CUH-P (Fig. 5g). For instance, *1p/19q* CODEL was found to be characterized by round nuclei with perinuclear halos, fried-egg-like tumor cells, calcification, and chicken-wire vasculature. Notably, beyond confirming known features, CAMPaS also highlighted less-recognized patterns, including the coexistence of spindle and plump spindle cells and high-grade histological features in *IDH*-wildtype, and the combination of perivascular lymphatic cuff and gemistocytic cells in *IDH*-mutant, offering visual cues for molecular prediction using WSIs in clinical workflows.

Lastly, in CUH-P, for *IDH* mutation (accuracy 84.5%, error case 55), *1p/19q* CODEL (accuracy 81.3%, error case 15), and *CDKN2A/B* HOMDEL (accuracy 63.3%, error case 44), pathologists identified representative histological confounders with cases that were both misclassified and ranked among the top 5% most ambiguous in the CAMPaS-learned feature space (Fig. 5h). For example, normal cerebral cortical gray matter with abundant neurons led to misclassification of *IDH* mutation, possibly because large neurons with prominent nucleoli tend to be misinterpreted as tumor cells by AI. Similarly, blood clot, appearing as necrosis-like uniformly red regions, was identified as a histological confounder for *CDKN2A/B* HOMDEL, the indicator of high-grade tumors in IDH-mutated glioma. Such findings can guide further improvements to CAMPaS using a pathologist-in-the-loop approach, enhancing its clinical reliability and interpretability.

#### 2.6.2 CAMPaS showed prognostic value

We validated the prognostic value of CAMPaS using both AI-predicted grades and continuous malignancy scores for both overall survival (OS) and progression-free survival (PFS) (Fig. 6a-g). For example, in TCGA (n=940), Kaplan-Meier (KM) curves restricted to 5-year follow-up demonstrated that the predicted Grade 4 (median PFS: 351 days) had significantly poorer survival than predicted Grade 3 (median PFS: 840 days) and predicted Grade 2 (median PFS: 1,197 days) (pairwise log-rank tests, all *P*<0.050; Fig. 6b), suggesting the prognostic value of AI-predicted grading.

**Fig 6.**
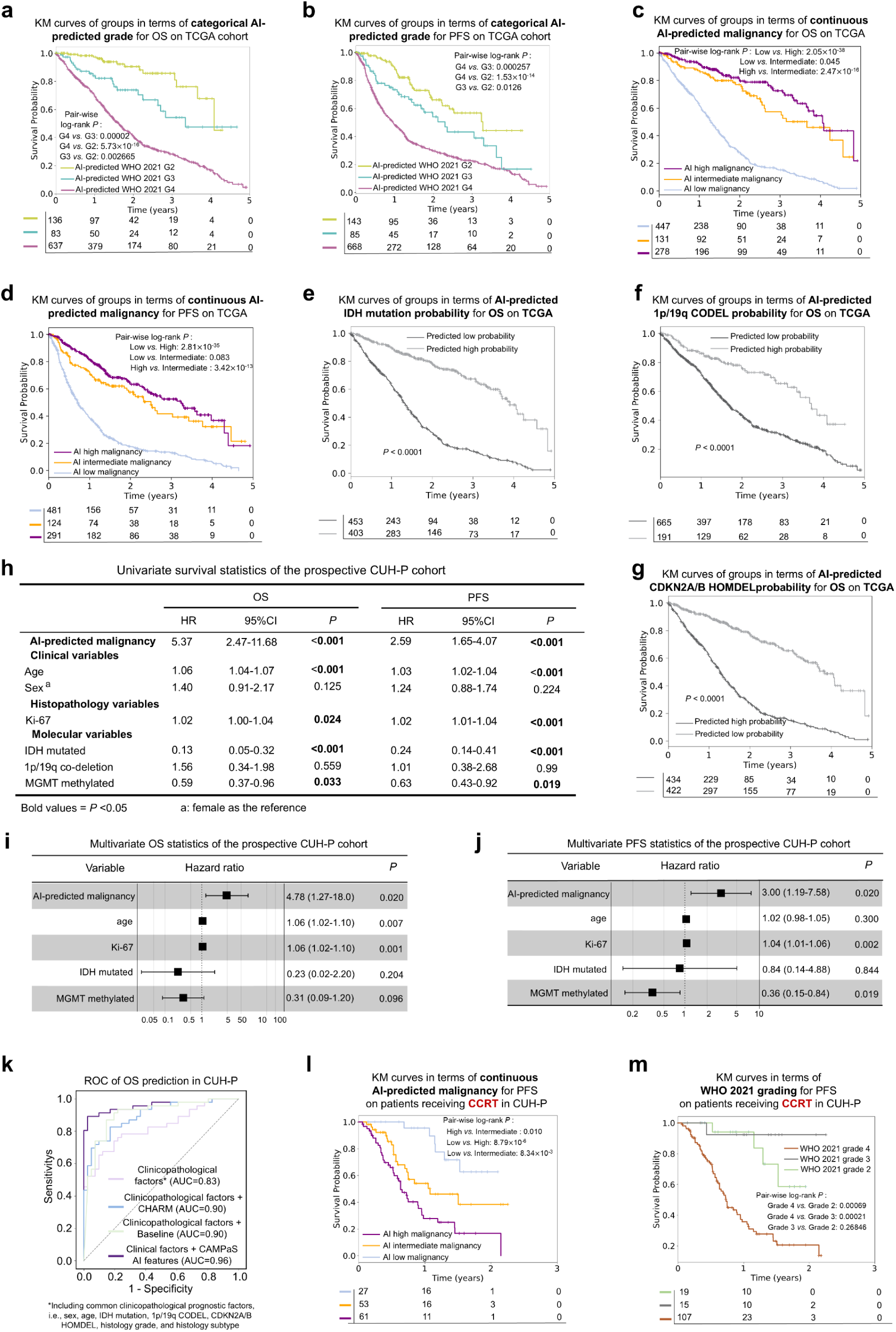
Clinical utility of CAMPaS in prognostic evaluation. **(a-b),** Survival analysis based on AI-predicted WHO 2021 histology grades. Kaplan-Meier (KM) curves show groups stratified by AI-predicted grades (G2: grade 2, G3: grade 3, G4: grade 4) in the TCGA cohort show significant differences in overall survival (OS) (**a**) and progression-free survival (PFS) (**b**), validating the prognostic value of AI-predicted grading. **(c-d),** Prognostic value of AI-predicted malignancy scores in the TCGA cohort. KM survival curves stratified by AI-predicted malignancy scores (low, intermediate, high) show significant differences in both (**c**) OS and (**d**) PFS. Pair-wise p-values of the log-rank test are shown. (**e-g**) KM survival curves show prognostic value of AI-predicted probability of the molecular markers, including *IDH* mutation (**e**), *1p/19q* CODEL (**f**) and *CDKN2A/B* HOMDEL (**g**) in the TCGA cohort. (**h-j**) Univariate and multivariate models including AI-predicted malignancy score and available established clinical, histopathological and molecular prognostic factors in the prospective CUH-P cohort. **h**, Univariate Cox regression analysis of OS (left) and PFS (right). AI-predicted malignancy scores show strong and significant associations with both OS (HR = 2.62, *P* = 0.003) and PFS (HR = 3.84, *P* < 0.001). (**i-j**) Multivariate Cox regression analyses further confirm that AI-predicted malignancy remains an independent prognostic factor for both OS (**i**, HR = 2.62, *P* = 0.003) and PFS (**j**, HR = 2.34, *P* = 0.038), after adjusting for known prognostic variables such as age, *Ki-67*, *IDH* mutation, and *MGMT* promoter methylation, which are all significant in univariate Cox regression analysis. **k,** The AI importance maps from CAMPaS achieve a higher AUC in OS prediction compared to those from CHARM or the CNN-based baseline, when integrating with clinicopathological factors, validated in the prospective CUH-P cohort. **(l-m),** KM analysis of AI-predicted malignancy scores (**l**) versus WHO 2021 grades (**m**) in patients treated with concurrent chemoradiotherapy (CCRT) in CUH-P cohort. Pairwise log-rank P-values show that AI-predicted malignancy scores significantly distinguish three risk groups on PFS modelling, better stratifying patients over the WHO 2021 grading, particularly for Grades 2 and 3.

We further stratified patients into low-, intermediate-, and high-risk groups based on AI-predicted malignancy scores. KM analyses showed that the three groups had significantly different OS and PFS (pairwise log-rank tests, all *P*<0.050; Fig. 6c-d).

These survival trends remained robust across molecular markers (Fig. 6e-g; Fig. S16), confirming the prognostic value of the malignancy scores.

In CUH-P, univariate Cox regression identified AI-predicted malignancy scores as a strong predictor of survival (OS: hazard ratio [HR] 5.37, 95% CI: 2.47-11.68, *P*<0.001; PFS: HR 2.59, 95% CI: 1.65-4.07, *P*<0.001; Fig. 6h). In multivariate models adjusted for clinical variables, the malignancy score retained independent significance (OS: HR=4.78, 95% CI=1.27-18.0, *P*=0.020; PFS: HR=3.00, 95% CI=1.19-7.58, *P*=0.020; Fig. 6i-j).

In CUH-P, integrating AI features with clinicopathological factors (AUC 0.96) significantly improved OS prediction compared to models only using clinicopathological factors (AUC 0.83; Fig. 6k). CAMPaS-derived features achieve better performance over CHARM-derived (AUC 0.90) and baseline-derived (AUC 0.90) features. Similar improvements were observed in TCGA, increasing AUC from 0.79 to 0.90 (Fig. S17).

Finally, we evaluated the prognostic value of CAMPaS in a subset (n=141) of CUH-P patients treated with standard concurrent chemoradiotherapy (CCRT) with temozolomide. KM curves showed significantly worse survivals in the AI-predicted high-malignancy group compared to intermediate- and low-malignancy groups (pairwise log-rank tests, all *P*<0.050; Fig. 6l). In contrast, histology-based and WHO 2021 integrated gradings showed similar trends but lacked statistical significance in stratifying Grades 2 and 3 (Fig. S18; Fig. 6m-n). When comparing the AI stratified subgroups with the WHO 2021 grades, we observed that among the WHO 2021 grade 4 patients (n=107; median PFS 267 days), CAMPaS classified six patients as low-malignancy (median PFS 420 days) and 40 patients as intermediate-malignancy (median PFS 274 days). CAMPaS may have achieved better stratification by utilizing more comprehensive features from the global slides via the multi-instance learning and mitigating heterogeneity via the attention mechanism. Together, these results indicate that CAMPaS shows potential for guiding personalized clinical decision-making.

## Discussion

The landscape of cancer diagnostics has transitioned from relying solely on histological features to integrating molecular markers. While this integration enhances diagnostic accuracy and biological relevance, it introduces practical challenges. To develop our previously benchmarking-validated research prototype into a robust, clinically usable diagnostic support tool, we proposed CAMPaS, a trustworthy AI system with tailored designs to capture intrinsic relationships among molecular markers and histology-molecular marker correlations, and enhanced architecture to handle real-world challenges such as dataset heterogeneity and incomplete molecular information.

The model performance benefits from being trained and validated on 3,367 patients (6,043 WSIs), achieving robust performance and interpretability in jointly predicting histology grading/subtyping, molecular markers, and integrated diagnosis. CAMPaS obtained an AUC of 0.946 and 0.955 in two prospective cohorts for integrated glioma diagnosis, comparable to state-of-the-art AI diagnostic tools in other cancers, such as breast cancer^44^ (AUC 0.943) and endometrial cancer^45^ (AUC 0.96). CAMPaS demonstrated robust performance across different numbers of WSIs per patient and varying training cohort sizes. It also performed consistently across patient subgroups and section modalities (FFPE and frozen), supporting its use in intraoperative settings where rapid diagnosis with frozen samples is needed. Moreover, CAMPaS remained reliable even as the proportion of missing molecular markers increased during training.

Few studies have developed models for glioma diagnosis, aligning with the WHO 2021 criteria. Despite success, most existing attempts were limited by small sample sizes and narrow scopes. Studies like DeepGlioma^10^, involving only ∼200 samples, need further robust validation. CHARM^11^ did not include key subtypes, such as Grade 4 astrocytoma with *CDKN2A/B* HOMDEL. Similarly, the ResNet^20^-based method^9^ may not fully comply with the WHO 2021 criteria, e.g., misclassifying GBM by applying redundant criteria other than IDH wildtype. Other studies^12,46^ were validated using a single cohort or lacked molecular-histological interpretability, thereby limiting their trustworthiness.

Our interpretability analyses provide insights into understanding glioma. Key molecular markers and histological features were consistently linked to high AI-predicted malignancy, reinforcing the model’s pathological grounding. Spatial transcriptomics further revealed strong spatial alignment between AI importance maps for predicting molecular markers and histology grades/subtypes. These findings highlight the model’s ability to capture meaningful cross-modal relationships.

Interestingly, genomic alterations in several tumor-related genes, such as *ATRX* and *OLIG2*, which are downstream of or frequently co-occur with *IDH* mutations, showed strong associations with our AI-predicted malignancy, despite not being included in the training. This may reflect the potential of our molecular graph modeling in capturing intrinsic relationships among molecular markers and predicting more candidate markers based on histology. Further, accurately predicting the additional diagnostic markers (including *TP53*, *PTEN*, and *EGFR*) by the pretrained CAMPaS demonstrated its potential for future versions of WHO criteria with expanding marker panels.

CAMPaS demonstrates strong clinical utility. Prospective validation illustrated that AI-generated visualizations significantly improved diagnostic accuracy and reduced discrepancies across pathologists of varying experience. Moreover, AI importance maps showed utilities in aiding pathologists to identify histological correlates of molecular markers, facilitating integrated diagnostics. This capability is particularly beneficial in time-sensitive clinical settings such as rapid intraoperative decision-making.

CAMPaS provides prognostic value. AI-predicted malignancy scores successfully stratified patients across cohorts, independently predicting survival and associated with transcriptional and immune cell profiles. This underscores the model’s transferability to downstream prognostic and biological inference tasks. In the subgroup receiving concurrent chemoradiotherapy, AI can better stratify patients over conventional metrics, highlighting its potential for personalized therapeutic guidance. Further, our results showed that high CAMPaS malignancy was associated with *CD38* expression, a chimeric antigen receptor T cell target^47^, which suggests its potential for selecting patients in new treatment development and warrants further investigation.

Given its cross-modal architecture and strong out-of-distribution performance, CAMPaS could serve as a pretrained backbone for transfer learning in other molecularly defined cancers, including endometrial cancer^48^ and renal neoplasia^49^. The modular design of CAMPaS, integrating vision transformers, molecular graphs, and uncertainty-aware multi-instance learning, supports plug-and-play adaptation to other diagnostic workflows beyond glioma. Future work will extend CAMPaS to other tumor types, potentially establishing a foundation model paradigm for computational molecular pathology.

Limitations include our focus on defined WHO diagnostic markers rather than broader panels. Future studies will expand markers in fine-tuning. In addition, some morphological correlations observed (e.g., structural changes) were not quantified due to the lack of labelled datasets. Finally, pathologists could further improve the AI model by pinpointing misinterpreted histological features in future development using a human-in-the-loop approach, while continuous prospective validation, in light of the evolving cancer therapeutics, remains essential for patient stratification.

In conclusion, CAMPaS offers robust AI modelling for integrated cancer diagnosis, trained on the largest glioma cohorts to date. It addresses key real-world challenges through advanced cross-modal AI methodology. It demonstrates clinical utility through improved diagnostic accuracy, interpretability, and prognostic stratification.

## Data Availability

All data produced in the present study are available upon reasonable request to the authors after peer-reviewing period

## Methods

### 1. Ethics and data privacy

The TCGA (phs000178), CPTAC (phs001287), GBMatch (EK078-2004, EK550/2005, EK1412/2014, EK 27-147/2015) and IvyGAP (PRJNA420741) study protocols were approved by the Ethical Committee at their participating centers. Protocols for the four in-house cohorts (i.e., TTH, SYSUCC, CUH-R, and CUH-P) were approved by the Ethics Committee of Beijing Tiantan Hospital (KY2023-160-02), Sun Yat-sen University Cancer Center (SYSUCC IRB: SL-A2021-053-02, SL-A2021-053-08, and SL-A2021-053-15), and Cambridge University Hospital (PRaM-GBM: CRUK/A19732; SIND: NCT04007185). For those enrolled in the prospective study, written consent was obtained, while for the retrospective study, a waiver of consent was signed when applicable. All research adhered to ethical standards, including the Declaration of Helsinki. Data from in-house cohorts were processed separately at each participating hospital.

### 2. Cohorts

All eligible patients from the participating centers within the given period were included. For retrospective cohorts, the TTH cohort recruited patients between February 2015 and January 2018. The CUH-R cohort included patients from January 2019 to October 2021. Cases were excluded based on the following criteria (Fig. S1): (1) without available WSls, (2) without any molecular markers of *IDH* mutation, *1p/19q* CODEL and *CDKN2A/B* HOMDEL, (3) WSIs with low image quality reviewed by our expert pathologist, i.e., no tumor tissue, poor tissue quality, or out-of-focus scanning. For prospective cohorts (i.e., CUH-P and SYSUCC), further exclusion criteria included: (1) suspicious histology of midline glioma, and (2) missing treatment and survival information due to referral to other hospitals. The CUH-P cohort involved patients recruited from November 2021 to February 2024, following the implementation of the WHO 2021 criteria. The SYSUCC cohort involved patients recruited between April 2022 and April 2024, with all participants diagnosed under WHO 2021 criteria. Of note, within trial frameworks in both CUH and SYSUCC, we recruited all eligible patients and included pathology analysis routinely implemented under WHO 2021 criteria. All prospective patients underwent surgical resection followed by adjuvant treatment including concurrent chemoradiotherapy (CCRT), short-course radiotherapy (SCRT), chemotherapy, or palliative care, where appropriate.

For the public cohorts, the TCGA included 1,122 patients with diffuse gliomas diagnosed from 1989 to 2013, mostly from the United States, with smaller groups from Sweden, Italy, Canada, Brazil, Germany, and Australia. Most patients had tumor resection followed by radiotherapy and chemotherapy. The CPTAC cohort recruited 99 glioma patients between 2016 and 2021, primarily from Russia, China, Poland, and the United States, with some from the Philippines and Bulgaria. These patients underwent tumor resection and received various treatments, including radiotherapy, chemotherapy, molecular therapy, and immunotherapy. The IvyGAP cohort included 41 patients recruited between 2006 and 2013, while the GBMatch cohort recruited patients between 2001 and 2016; all underwent tumor resection.

### 3. Datasets

We included patients with diffuse gliomas and collected frozen and formalin-fixed paraffin-embedded (FFPE) tumor sections, along with clinicopathological data and molecular test results from four public cohorts and four in-house clinical cohorts (Table S1). For all cohorts, at least one H&E-stained slide of the glioma surgery-derived specimen was included for each patient, depending on availability. For the in-house cohorts, we collected one diagnostic H&E-stained slide per patient, each from an FFPE tissue block. H&E slides were scanned at x40 magnification using Leica Aperio scanner AT-2 (resolution 0.2468 μm per pixel), EasyScan scanner SZ-20 (resolution 0.2650 μm per pixel) and Nanozoomer scanner s60 (resolution 0.2521 μm per pixel), for CUH, SYSUCC and TTH, respectively.

Following the aforementioned criteria, 3,367 patients (6,043 WSIs) were included: 940 (2,633 WSIs) from TCGA, 81 (192 WSIs) from CPTAC, 218 (370 WSIs) from GBMatch, 33 (753 WSIs) from IvyGAP, 1,115 (1,115 WSIs) from TTH, 422 (422 WSIs) from CUH-R, 354 (354 WSIs) from CUH-P, and 204 (204 WSIs) from SYSUCC.

More details of molecular and survival data can be found in Supplementary section 2.

### 4. Model performance evaluation

Threefold cross-validation was performed on the 2,633 WSIs reserved for supervised training, and the best-performing architecture and hyperparameters were selected based on the highest mean AUC across the folds. A receiver operating characteristic (ROC) curve analysis and an AUC analysis were done with 95% Wald confidence interval (CI) using the continuous AI-predicted malignancy score with bootstrapping (300 sample size and 50 sample times).

For further evaluation, six additional metrics with 95% confidence intervals were reported: accuracy, sensitivity, specificity, precision, recall, and F1-score. Notably, for all comparison methods, including CHARM^11^, CLAM^24^, TransMIL^25^, and the CNN-based baseline^9^, separate models were trained independently for histology grading/subtyping, molecular marker prediction, and integrated glioma diagnosis, as these methods are not designed to jointly handle multiple tasks.

Finally, to validate model robustness across varied WSIs, we used the most representative slide as the baseline, defined as the slide with the lowest prediction uncertainty (i.e., the lowest Shannon entropy among a patient’s WSIs).

### 5. Cross-modal AI diagnostic model

#### 5.1 Framework

As shown in Extended Fig. 3, given the extracted representation features 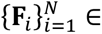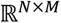 (with N = 2,500 patches and M = 1,024 for feature dimension) from the cropped patches 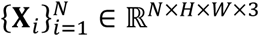 (with height H = 224 pixel, width W = 224 pixel and 3 denotes the number of RGB channels), CAMPaS predicted: (1) Molecular markers, including *IDH* mutation (𝑙^_idh_ ∈ ℝ^2^), *1p/19q* CODEL ( 𝑙^_1p/19𝑞_ ∈ ℝ^2^ ) and *CDKN2A/B* HOMDEL ( 𝑙^_cdkn_ ∈ ℝ^2^ ); (2) Histology grading ( 𝑙^*_grading_* ∈ ℝ^3^ ) and subtyping ( 𝑙^*_subtyping_* ∈ ℝ^3^ ); (3) Final integrated diagnosis of glioma (𝑙^*_glio_* ∈ ℝ^6^). CAMPaS adopted a hierarchical multi-task learning framework based on a top-down vision transformer backbone. For translational efficacy, in this paper, we specially designed a bi-directional attention module and a noise-robust learning module to solve the real-world challenges of dataset heterogeneity and partially missing molecular markers, respectively.

#### 5.2 Bi-directional attention module

The bi-directional attention module was designed to extract robust features from WSIs for predicting molecular markers as well as for histology grading and subtyping (Extended Fig. 3; Fig. S19). To mitigate dataset heterogeneity, this module dynamically identified critical WSI regions by applying thresholds to AI-generated importance maps. These selected regions then guided the reweighting of WSI patches, thereby suppressing redundant or noisy areas.

Specifically, the input patch-level features 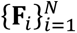 were first processed through three cascaded transformer blocks to effectively extract WSI features. After an initial warm-up training stage (lasting for several epochs, i.e., 10 in this paper), the bi-directional attention mechanism was repeatedly applied throughout the remaining training phases. Within each iteration of the bi-directional attention guidance, AI importance maps highlighting the most discriminative regions for each target class were generated (See Section 8 in Methods). These maps subsequently served as attention masks, directly multiplied with the features 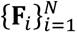 in the subsequent epochs. By doing so, the model progressively focused more precisely on class-specific informative regions for downstream tasks, while simultaneously suppressing noisy regions arising from WSI heterogeneity. For instance, in the histology grading task, dominant features indicative of tumor grading were prioritized, enabling the model to explicitly filter out distracting features within the WSIs. Details of generating the binarized attention guidance are elaborated as follows.

Taking the histology grading task as an example, we employed a channel-adaptive threshold truncation strategy on the AI importance maps of grades 2, 3, and 4, i.e., 𝑆_𝐺2_, 𝑆_𝐺3_, 𝑆_𝐺4_. For each channel 𝑐 of the three grades, we computed the mean saliency conditioned on the network’s predicted class 𝑦_pred_:

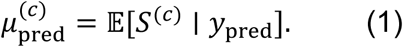

Pixels in the raw importance map 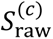 of each grade exceeding this threshold were retained, whereas those below were set to the channel’s minimum saliency 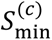:

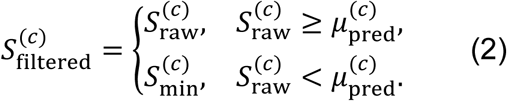

A subsequent linear normalization projected the truncated map onto the unit interval:

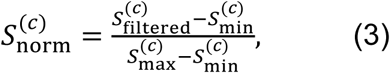

where 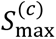 denotes the channel’s maximum saliency. This step simultaneously removed weak responses and unified saliency scales.

We then defined a fusion weight function based on the predicted grading category 𝑐, leveraging indicator functions 𝕀(⋅) to assign full attention to the predicted class and partial attention to non-predicted classes:

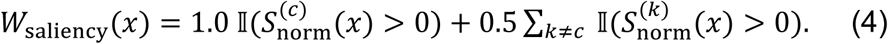

Finally, the fused weight map 𝑊_saliency_ was applied to the initial feature map 𝐹_init_ via the Hadamard product to produce the binarized attention guidance 𝐹_attn_, output of the histology grading task:

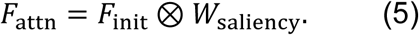

Similarly, the binarized attention guidance for the histology subtyping task can be derived.

Notably, the attention module incorporated a unified adaptive color-space normalization strategy^50^ to mitigate domain shifts across datasets. This normalization dynamically adjusted the color distribution of input WSIs to align with a reference domain, thereby ensuring consistent visual features across varying staining protocols and scanners.

### 5.3 Noise-robust learning module

Building on the robust features extracted by the bi-directional attention module, we introduced a novel noise-robust learning module to address label uncertainty caused by incomplete molecular annotations in real-world datasets. Within this framework, we incorporated the molecular graph subnet, histology prediction subnet, and cross-modal interaction subnet from our previous work^13,14^ to support molecular marker prediction, histology grading/subtyping, and modeling histology-molecular relationships (Figs. S20-21). Compared to our previous designs, we further enhanced the cross-modal interaction subnet, as introduced in Supplementary section 7.

In the noise-aware training process, we first estimated the uncertainty level of each WSI based on the availability of its molecular marker labels, achieved through entropy-based quantification (including two steps of consistency-based categorization, and entropy-guided noise assessment), where higher entropy, showing greater ambiguity or missing annotations, indicates a higher likelihood of label noise. Based on these estimates, we categorized WSIs as either noisy or clean. Noisy samples were applied to an adaptive loss modulation strategy to down-weight the impact of unreliable labels, thereby reducing the risk of overfitting to noise. In contrast, clean samples were trained using a standard cross-entropy loss to provide accurate supervision. This overall procedure formed an uncertainty-adaptive learning framework, dynamically calibrating the learning signal according to the estimated label quality (Details in Supplementary Section 8). Model implementation details can be found in Supplementary section 9.

### 6. Model training and validation strategy

We trained and assessed our model in three stages as follows.

***Stage I: Training***. To empower the AI model for effective cross-modal modeling under the WHO 2021 criteria, we first trained our model on TCGA, a large-scale multicenter cohort with both molecular markers and histology grading.

***Stage II: Fine-tuning.*** To enhance the generalizability of our model on heterogeneous datasets, we fine-tuned and tested our model on five retrospective cohorts, which demonstrate heterogeneity in color and cellular distributions. In model fine-tuning, we included all samples meeting the general inclusion and exclusion criteria.

***Stage III: Testing.*** To further evaluate the model in real-world clinical scenarios, we recruited two prospective cohorts diagnosed under the WHO 2021 criteria.

### 7. Association with multimodal diagnostic factors analysis

For analyzing the association of AI-predicted malignancy scores with diagnostic factors (i.e., clinicopathological factors and molecular markers), patients were first clustered into three subgroups according to their AI-predicted malignancy scores with agglomerative clustering^28^. This unsupervised approach iteratively merged with the closest clusters based on the score similarity, minimizing within-cluster variance. We determined the optimal number of clusters (k = 3) based on the dendrogram structure and interpretability in the clinical context, resulting in low, intermediate, and high malignancy groups.

Then, integrated visualization of AI-predicted malignancy scores and related clinical, molecular, and histopathological features (for TCGA, SYSUCC, and CUH-P cohorts) was performed (Fig. 3a and Fig. S7). We further performed multiple single linear regression analyses using the continuous malignancy score as a dependent variable and the clinicopathological data as the regressor. Statistical tests were two-sided, with statistical significance accepted if *P* values < 0.050. Of note, in Fig. S7, the potential diagnostic molecular markers (both CNV- and SNP-level) were selected as the markers with at least 5% alteration proportion from the cBioPortal.

### 8. Generating AI importance maps under MIL settings

To obtain the AI importance map, following a well-recognized MIL-based network visualization method^51^, we utilized the mechanisms of the class activation map (CAM) method^52^ and extended it to the MIL settings for visualizing the AI decisions for tasks of histology grading, subtyping, and molecular markers prediction. Following previous work^13,14^, for WSIs with fewer than 2,500 original patches, we averaged the importance scores of the 2,500 patches assigned to each original patch. For WSIs with more than 2,500 original patches, we applied bilinear interpolation to align the 2,500 importance scores with the original patch grid, ensuring accurate and consistent visualization.

### 9. AI-predicted malignancy score

Based on the AI prediction of molecular markers and histology grades/subtypes, the AI-predicted malignancy score indicated the probability of being high grade, including Grade 4 GBM and Grade 4 Astrocytoma in WHO 2021 criteria. Formally, the malignancy score, 𝑙^_malig_ ∈ ℝ^1^) was defined as follows:

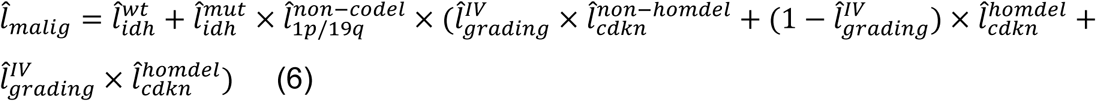

In the above equation, 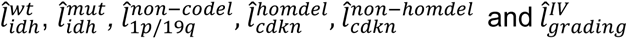 indicated the AI predicted probability of *IDH* wildtype, *IDH* mutated, *1p/19q* CODEL, *CDKN2A/B* HOMDEL, *CDKN2A/B* non-HOMDEL, and histology grade 4, respectively.

### 10. Morphological interpretability analysis

To investigate morphological correlations between AI importance maps and histopathological metrics, AI importance maps generated by the AI model for histology grading were analyzed. Specifically, the PCC was calculated between the spatially averaged AI importance maps and 50 commonly utilized histopathological metrics, as annotated by expert pathologists in the GBMatch cohort (Fig. 3d, Fig. S9). To rigorously evaluate the interpretability and precision of our model, we compared its performance with CHARM (ViT-based) and a CNN-based model. Notably, in subsequent association analyses, a subset of the most important AI features was used, specifically the top K% (K = 5, 25, or 50) elements of the layer prior to the MLP layers, selected according to their values.

To further validate these morphological insights, we quantitatively assessed neuropathologist-annotated histological features, particularly the percentage of tumor nuclei and necrosis area, against AI-defined malignancy subtypes in the TCGA cohort (Fig. 3e-f).

### 11. Bulk-level and spatially resolved transcriptomic correlation analysis

#### Bulk gene expression-level association

To explore bulk-level transcriptomic correlates of AI’s decision on molecular markers, we performed comprehensive analyses integrating AI importance maps with gene expression data (normalized with Transcripts Per Million (TPM)) from the TCGA cohort. Initially, bulk-level transcriptomic associations were evaluated by calculating PCC between the spatially averaged AI importance maps and expression levels of selected brain tumor-related and immune-related genes. Gene sets utilized for these analyses included 140 brain tumor-associated genes (including potential diagnostic marker genes in WHO 2021^3^, and transcriptomics-level GBM subtyping marker genes^31^) and top 100 highly variable immune-related genes derived from established databases, such as ImmPort^32^ (Fig. 3f-g; Figs. S11-12). Gene-wise correlation significance was assessed using the PCC metric, and statistical significance was established using a threshold of *P* < 0.05.

#### Immune cells-level association

A novel cell-type deconvolution algorithm, i.e., BayesPrism^37^, was used for estimating the relative proportions of 9 common glioma immune cell types in TCGA cohort based on patients’ bulk transcriptomics data, using single-cell RNA sequencing data^36^ as the deconvolution reference. Then, we used Ward linkage-based hierarchical clustering method^53^ to generate the heatmap of the cell-type proportions, ordered by three clusters representing the three subtypes in terms of AI-predicted malignancy. Further, the association between the AI-predicted malignancy scores and the estimated cell-type proportions was performed using the log2 (transformed proportion of the immune cell subset) as a fraction of the whole tumor, using the leukocyte fraction values. Linear regressions were performed with the CAMPaS continuous risk scores as the independent variable. Two-sided P values <0.050 are accepted as significant.

Differentially expressed genes were assessed between high-risk and low-risk cases stratified by predicted AI malignancy score, using the Scipy Python package (v.1.11.1) and statsmodels Python package (v.0.14.1). Genes with a likelihood ratio test *P* value were accepted if *P* <0.050.

#### Spatial transcriptomics-level association

For spatially resolved transcriptomic correlation, we utilized glioma spatial transcriptomic (ST) profiling data, along with spatially aligned H&E-stained WSIs, from a published work^40^, generated using the 10X Visium^54^ platform. Raw data preprocessing included normalization, scaling, and dimensionality reduction to produce spatial gene expression profiles. The paired WSIs were preprocessed for AI prediction following the same procedures used for other cohorts as described above. Decision-relevant spatial regions (for molecular marker prediction) identified by AI from WSI were then compared to these spatial gene expression patterns (Fig. 4d). Specifically, we computed cosine similarity scores between the AI importance maps and spatial expression patterns of key tumor-related and immune-related genes, thereby quantitatively assessing the spatial coherence between AI predictions and gene expression profiles.

### 12. Clinical utility in diagnosis

To prospectively validate the clinical utility of CAMPaS in glioma diagnostics, a structured two-stage evaluation was conducted across two independent cohorts: CUH-P (n = 354) and SYSUCC (n = 203). The validation incorporated neuropathologists categorized into junior, intermediate, and senior levels, with clinical experience in neuropathology of 1 (C.L.), 8 (X.Z.), and 15 (W.H.) years. These pathologists were first trained for using the website of CAMPaS and then conducted the two-stage evaluation based on the H&E sections from tumor samples routinely obtained via surgery. Details of the two-stage evaluation process can be found in Supplementary section 10.

Pathologists’ diagnostic performance before and after AI assistance was evaluated based on the final WHO diagnosis confirmed after molecular testing as the reference standard. Specifically, the diagnostic performance was assessed by classification accuracy and ROC analysis across neuropathologists with different levels of experience. Confidence intervals (CIs) for accuracy estimates were calculated using a 95% CI to robustly evaluate improvements associated with AI assistance.

Furthermore, to elucidate the histological features driving diagnostic improvements with AI assistance, the top 5% of WSI regions in terms of heatmap pixel values in the corresponding AI importance maps were extracted. Then, these selected regions were reviewed separately by neuropathologists across all levels to characterize and summarize the representative morphological patterns associated with AI-guided diagnostic gains.

### 13. CAMPaS usability analysis

We evaluated the usability of the website of CAMPaS across pathologists (n=10) in the department of SYSUCC, using system usability scale (SUS) score of ten standard evaluation questions (Table. S15). We first calculated the SUC scores for all neuropathologists and then averaged them for the final SUS score.

### 14. Prognostic efficacy analysis

The prognostic efficacy of AI-predicted malignancy features was systematically assessed through survival analyses in both the retrospective TCGA and prospective CUH-P cohorts. The prognostic significance was evaluated for OS and PFS using KM survival analysis^55^ and two-sided log-rank tests. In continuous AI malignancy survival analysis, cutoffs for the CAMPaS malignancy groups were defined by taking the tertiles of the distribution of CAMPaS predicted malignancy scores in the cohorts. Survival outcomes across these groups were compared and pairwise log-rank tests were conducted to establish significant differences among these groups.

Additionally, univariate and multivariate Cox proportional hazards regression analyses were performed to quantify the prognostic value of continuous malignancy scores and established clinical (sex and age), histopathological (*Ki-67* index), and molecular (*IDH* mutation, *1p/19q* CODEL and *MGMT* promotor methylation) variables. Hazard ratios (HRs), along with 95% CIs and corresponding *P* values, were calculated to determine the independent prognostic significance and effect size.

To evaluate the incremental prognostic value of AI-derived features, a logistic regression model taking the learned CAMPaS features and clinical diagnostic factors (including sex, age, histology grading and subtyping, *IDH* mutation *1p/19q* CODEL and *CDKN2A/B* HOMDEL status) was trained to predict median OS in both TCGA and CUH-P cohorts. The model was trained with a learning rate of 0.003 and a batch size of 4 on the same device as in training the CAMPaS. The compared models were logistic regression models taking only the clinical diagnostic factors, adding the learned CHARM features, or adding the learned baseline CNN-based features as input. Predictive performance was quantified using ROC analysis, with statistical confidence assessed using 95% CIs.

To evaluate the prognostic relevance of the features learned by CAMPaS, we computed malignancy scores for patients in the CUH-P who received standard CCRT (n =141).

PFS probabilities were modelled using the KM-based method. To assess treatment outcomes, patients were stratified into malignancy groups based on CAMPaS-predicted scores. Prognostic discrimination was compared against conventional histology-based grading, serving as a baseline reference.

For all log-rank tests, statistical significance was accepted for *P* < 0.050. Benjamini-Hochberg False Discovery Rate (FDR) correction was utilized for multiple comparisons. Software details can be found in Supplementary section 11.

## Data availability

The TCGA histology images, corresponding diagnostic labels, clinicopathological data, and gene expression data are available from NIH genomic data commons (https://portal.gdc.cancer.gov/). Other diagnostic molecular markers (both CNV- and SNP-level) for TCGA cohort are from cBioPortal (https://www.cbioportal.org/), while survival information and pathologists annotated necrosis and angiogenesis scores are from previous work^56,57^. The CPTAC histology images and corresponding diagnostic labels are available from the TCIA CPTAC Pathology Portal (https://cancerimagingarchive.net/datascope/cptac/) and cBioPortal, respectively. The IvyGAP histology images, pixel-wise annotations and corresponding diagnostic labels are from its website (https://glioblastoma.alleninstitute.org/). The GBMatch histology images, pathologists annotated histology metrics and corresponding diagnostic labels are available at its website (https://www.medical-epigenomics.org/papers/GBMatch/). Spatial transcriptomics data is available at Datadryad (https://datadryad.org/dataset/doi:10.5061/dryad.h70rxwdmj).

Restrictions apply to the availability of the raw in-house data, which was used with institutional permission through ethics approval for the current study, and are thus not publicly available. Please email all requests for academic use of raw and processed data to the corresponding author. All requests will be evaluated based on institutional and departmental policies to determine whether the data requested is subject to intellectual property or patient privacy obligations. Data can only be shared for non-commercial academic purposes and will require a formal material transfer agreement.

## Code availability

Codes are available at https://github.com/XiaofeiWang2018/CAMPaS.

## Acknowledgements

This work was supported by the NIHR Brain Injury MedTech Co-operative, the NIHR Cambridge Biomedical Research Centre (NIHR203312), Addenbrooke’s Charitable Trust, and the National Natural Science Foundation of China (grant numbers 62271475 and 82102877). This work presents independent research funded by the National Institute for Health and Care Research (NIHR). The views expressed are those of the author(s) and not necessarily those of the NHS, the NIHR or the Department of Health and Social Care. Xiaofei Wang reports financial support by China Scholarship Council. Stephen Price reports financial support by National Institute for Health and Care Research. Chao Li reports financial support by the Guarantors of Brain.

---

**Extended Fig 1.**
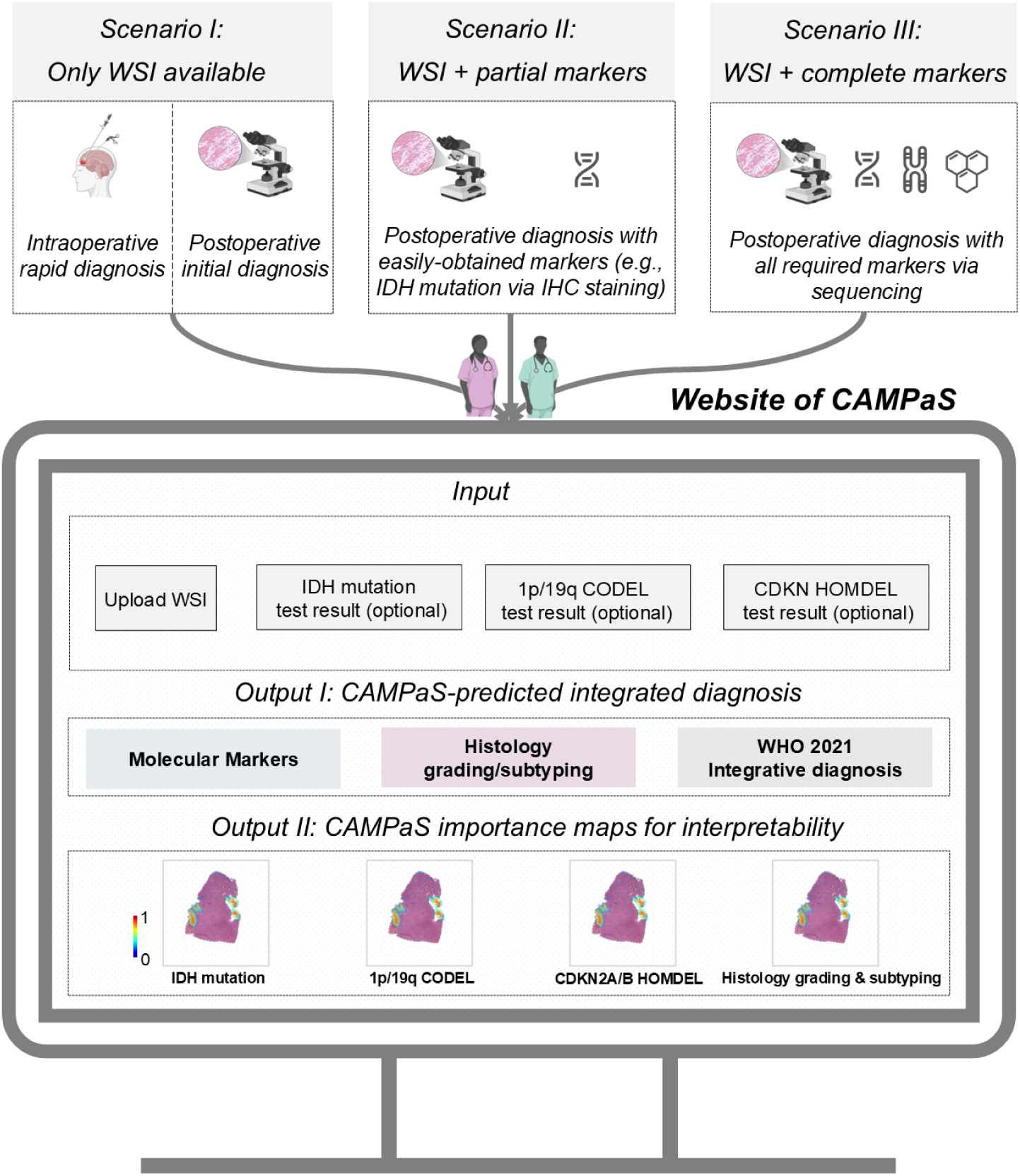
Interactive CAMPaS website supporting diverse clinical scenarios. The CAMPaS web platform is designed to support a broad range of real-world diagnostic workflows in glioma diagnosis, accommodating varying levels of available molecular information. It is tailored to the following scenarios: (**I**) WSI-only input: Typical for intraoperative rapid diagnosis during surgery or postoperative initial assessment using limited tissue and H&E staining only. (**II**) WSI + partial molecular markers: Applies to postoperative diagnosis when limited yet informative markers (e.g., *IDH* mutation) are available, often via immunohistochemistry (IHC). (**III**) WSI + complete molecular markers: Applies to postoperative diagnosis when all diagnostic markers (e.g., *IDH* mutation, *1p/19q* CODEL, and *CDKN2A/B* HOMDEL in WHO 2021 criteria) are available, yet no histology features of molecular markers are provided. For all scenarios, users can upload WSI and available markers through the web interface. The platform provides: (**Output I**) Integrated diagnostic predictions combining histology and molecular features under the WHO 2021 criteria. (**Output II**) AI-importance maps for interpretability, highlighting histopathological regions associated with each molecular marker and histology grade/subtype.

**Extended Fig 2.**
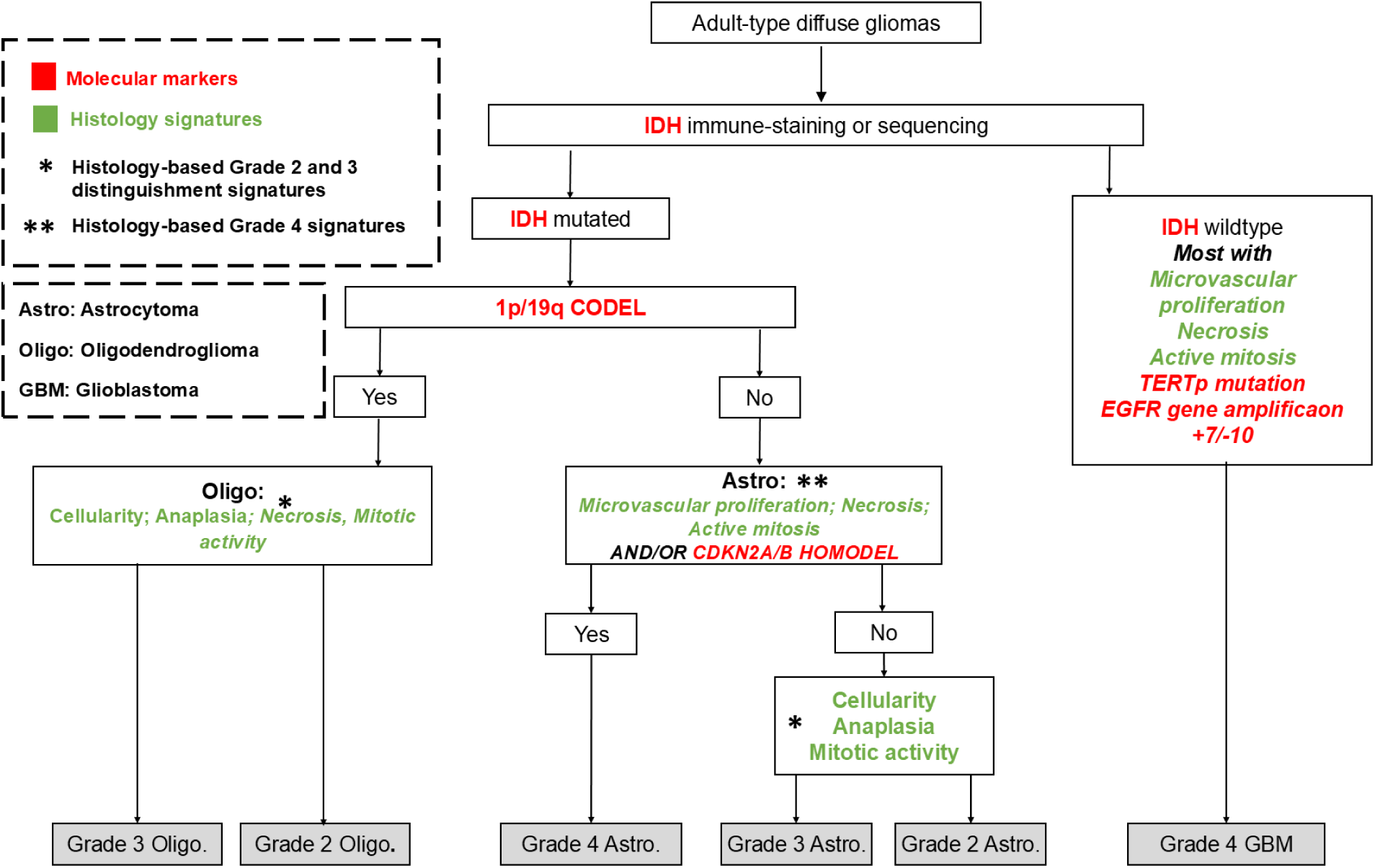
Integrated glioma diagnostic pipeline under WHO 2021 criteria. See Supplementary section 4 for the diagnostic scheme for both retrospective and prospective cohorts.

**Extended Fig 3.**
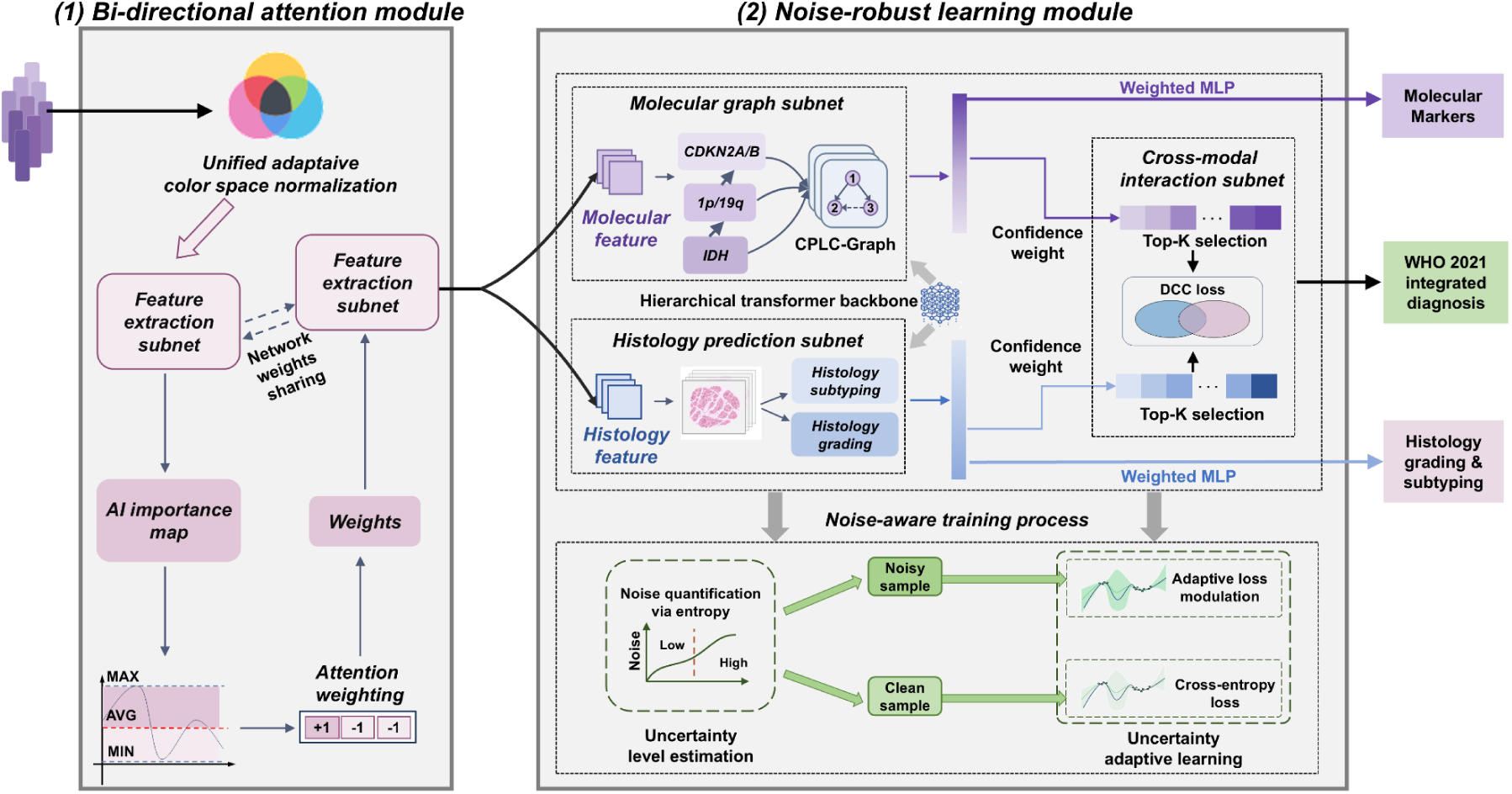
Illustration of the CAMPaS framework. Taking WSI patch features as input, CAMPaS outputs the predictions of molecular markers (i.e., *IDH* mutation, *1p/19q* CODEL and *CDKN2A/B* HOMDEL), histology grading (i.e., Grade 2, 3 and 4), and subtyping (i.e., Oligodendroglioma, Astrocytoma, and GBM), as well as the final WHO 2021 glioma diagnosis. Built on a hierarchical transformer backbone, CAMPaS integrates two key modules for clinical translation: a bi-directional attention module to model WSI heterogeneity and a noise-robust learning module to address label noise arising from missing molecular markers, as detailed below. **(1)** The bi-directional attention module is designed to extract robust WSI features for both molecular marker prediction and histology grading/subtyping. To address dataset heterogeneity, critical WSI regions are dynamically selected by thresholding the AI importance map. These regions are then used to reweight WSI patches, effectively filtering out redundant or noisy regions. **(2)** Based on the robust features extracted in the bi-directional attention module, we further propose a novel noise-robust learning module to mitigate the label uncertainty caused by partial molecular labels in real-world datasets. The key idea of this learning strategy is to first estimate the uncertainty level of each WSI based on the missing molecular markers and then perform an uncertainty adaptive learning process to account for the degree of label noise. Moreover, during this process, the molecular graph, histology prediction, and cross-modal interaction subnets from previous work^13,14^ are incorporated for predicting molecular markers, histology grading/subtyping, and modelling the histology-molecular interaction. In the molecular graph subnet, a Co-occurrence Probability and Label-Correlation Graph (CPLC-Graph)^13^ is designed to capture the intrinsic dependencies among molecular markers. Meanwhile, in the cross-modal interaction subnet, patch-level importance scores from both molecular and histology predictions (termed "confident weights") are subjected to a Top K (where K is a hyperparameter) selection, retaining the most reliable patches for further analysis. A Dynamic Confidence Constraint (DCC) loss is applied to guide multimodal interaction and reduce the impact of unreliable WSI regions. Finally, we propose a noise-aware training process. During model training, the first step involves estimating the uncertainty level of each WSI based on the availability of corresponding molecular labels. This is achieved through entropy-based quantification, where high entropy reflects greater ambiguity or missingness in molecular annotations, indicating a higher likelihood of label noise. WSIs are thus categorized into noisy or clean samples accordingly. For noisy samples, an adaptive loss modulation strategy is employed to down-weight the influence of unreliable labels, mitigating the risk of overfitting to noise. For clean samples, the model is trained using a standard cross-entropy loss, enabling accurate supervision. This process constitutes an uncertainty adaptive learning framework, which dynamically calibrates the learning signal based on estimated label quality. A detailed description of CAMPaS can be found in the Methods.

**Extended Fig 4.**
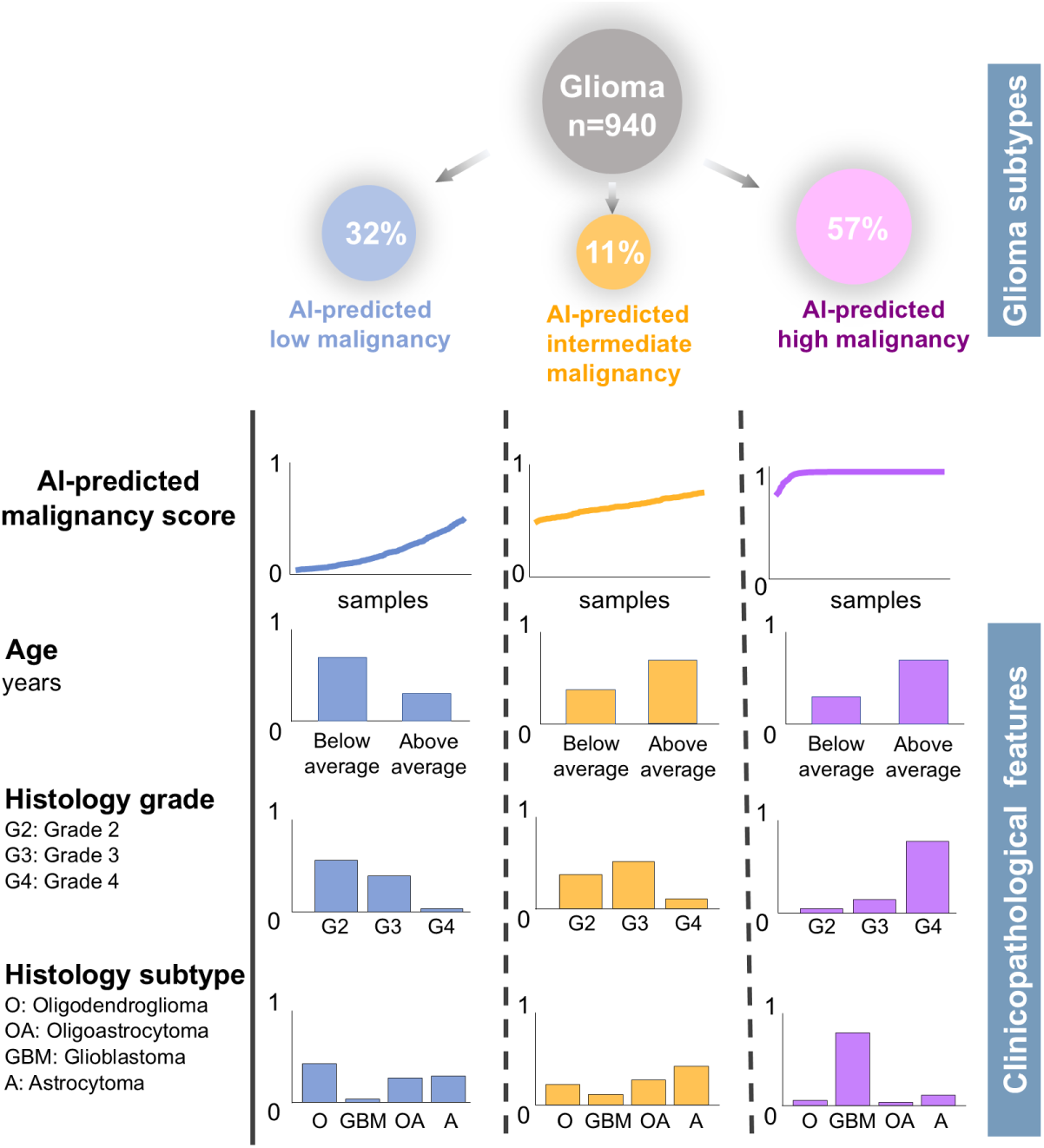
Overview of three subtypes stratified via AI-predicted malignancy scores. An integrative analysis of 940 gliomas in TCGA yields three subtypes with distinct clinicopathological characteristics. The three subtypes are identified by extracting the top-ranked features per subtype using AI-predicted malignancy scores with agglomerative hierarchical clustering^28^. The subtype of low AI-predicted malignancy (32%) is enriched for lower-grade tumors, predominantly astrocytoma, oligodendroglioma, and oligoastrocytoma, with more younger patients. The subtype with AI-predicted intermediate malignancy (11%) presents moderate AI-predicted malignancy scores and mixed histology grades. The subtype of AI-predicted high malignancy (57%) is dominated by glioblastoma, often in older patients, reflecting more aggressive disease. These trends suggest that AI-predicted malignancy scores effectively stratify glioma patients into clinically and biologically meaningful subgroups.

**Extended Fig 5.**
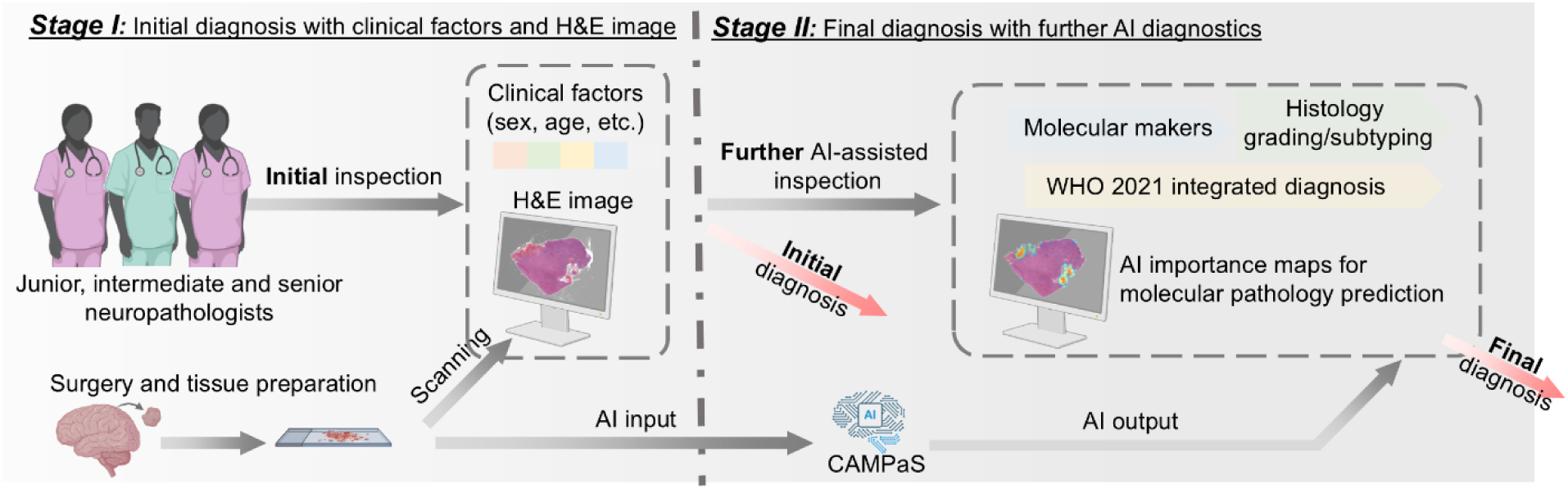
Schematic diagram of two-stage evaluation in assisting pathologists’ diagnosis. Prospective validation for clinical diagnosis comprises two stages and involves three neuropathologists with junior, intermediate, and senior levels of experience. In stage I, each pathologist diagnoses based on the WSI and clinicopathological factors, including age, sex and Ki-67 index. In stage II, the same randomized cases are re-evaluated with the addition of AI prediction results and corresponding AI importance maps.

## Footnotes

1 Patients with their glioma grading remained or updated, when transferring from previous diagnosis criteria to the current WHO 2021 scheme.

2 AI importance maps highlighted regions in WSIs relevant to predicting molecular markers and histology grading/subtyping, while AI-predicted malignancy scores reflected the likelihood of high-grade tumors based on WHO 2021 criteria.

3 The top 5% important features were further used for prognostic validation in Section 2.6.2

